# Plasma cfChIP-seq for non-invasive identification of autoimmune liver diseases

**DOI:** 10.1101/2023.06.04.23290776

**Authors:** Gavriel Fialkoff, Ami Ben Ya’acov, Israa Sharkia, Ronen Sadeh, Jenia Gutin, Abed Khalaileh, Ashraf Imam, Rifaat Safadi, Ofir Hay, Nofar Rosenberg, Chen Goldstein, Esther Harpenas, Yael Milgrom, Eithan Galun, Eyal Shteyer, Nir Friedman

## Abstract

**Background:** Autoimmune hepatitis (AIH) is a chronic immune-mediated liver disease associated with substantial morbidity and mortality. Diagnosis and monitoring typically rely on liver biopsy, despite its invasiveness and limitations. Liquid biopsies hold potential as non-invasive alternatives, but their utility in AIH remains underexplored.

**Objective:** To evaluate whether plasma-based chromatin immunoprecipitation followed by sequencing (cfChIP-seq) can detect hepatocyte-specific transcriptional activity in AIH and serve as a diagnostic and monitoring biomarker.

**Design:** We profiled circulating cell-free nucleosomes marked by H3K4me3—a histone modification marking active and poised promoters—in plasma samples from AIH patients (43 pediatric, 49 adult), healthy controls (6 pediatric, 1240 adults), and patients diagnosed with other liver diseases (15 pediatric, 175 adult).

**Results:** cfChIP-seq revealed immune-related hepatocyte transcriptional signatures representing intrahepatic activity unique to AIH patients. RNA-seq of matched liver biopsies further corroborates these findings. A score trained on cfChIP-seq profiles from AIH and metabolically associated steatohepatitis (MASH) patients discriminated liver autoimmune diseases (including AIH) from other liver conditions, including MASH and drug-induced liver injury (DILI), with high accuracy on independent validation cases (AUC = 0.94; 95% CI 0.84-1). Combined with a second cfChIP-seq-based classifier for AIH vs. PSC/PBC, we distinguish between AIH and biliary autoimmune diseases (AUC = 0.92; 95% CI 0.83-1).

**Conclusion:** Plasma cfChIP-seq captures hepatocyte disease-specific gene activity in AIH patients and offers a non-invasive, accurate method for diagnosing and monitoring AIH. This approach has the potential to reduce the reliance on liver biopsy, improve diagnostic precision, and provide novel insights into AIH pathogenesis.

**Key messages:** *What is already known on this topic:* Autoimmune hepatitis (AIH) is a chronic liver disease diagnosed and monitored primarily through liver biopsy, an invasive procedure with associated risks and sampling errors. Non-invasive alternatives are urgently needed but have shown limited disease specificity to date.

*What this study adds:* This study shows that cfChIP-seq,a plasma-based assay detecting gene activity in turned over cells including from liver, can accurately distinguish AIH from other liver conditions. The cfChIP-seq signal in AIH patients reflects intrahepatic immune activity and correlates with findings from liver biopsy.

*How this study might affect research, practice or policy:* By capturing disease-specific intrahepatic disease activity, cfChIP-seq provides positive indication of disease etiology. As such, cfChIP-seq has the potential to reduce the need for liver biopsies in AIH by providing a specific, non-invasive diagnostic tool. Its application in clinical practice could improve patient monitoring, support earlier intervention, and guide personalized management.

## Introduction

Patients presenting with persistently elevated liver transaminases represent a common and clinically challenging diagnostic scenario in hepatology. In many cases, non-invasive evaluation—including viral serology, metabolic assessment, drug exposure history, and imaging—fails to establish a definitive diagnosis. At this point, clinicians are often faced with the critical decision of whether to proceed with liver biopsy, an invasive procedure associated with pain, bleeding risk, sampling error, and limited suitability for repeated assessment.

A major indication for liver biopsy in this context is the need to exclude or confirm autoimmune hepatitis (AIH). AIH is a chronic immune-mediated liver disease that requires long-term immunosuppressive therapy, making accurate diagnosis essential. However, AIH frequently presents with non-specific clinical and biochemical features that overlap with other liver diseases, including metabolically associated steatohepatitis (MASH), drug-induced liver injury (DILI), viral hepatitis, and cholestatic disorders. As a result, liver biopsy is often performed not to confirm a clear clinical suspicion, but rather to resolve diagnostic uncertainty (“Autoimmune Hepatitis: A Comprehensive Review” 2013; Mieli-Vergani et al. 2018).

Despite the availability of diagnostic scoring systems and serological markers—such as autoantibodies and serum immunoglobulin G (IgG)—the diagnosis of AIH remains imperfect. Autoantibodies may be absent, particularly in pediatric patients and in acute or early disease, and IgG levels can be normal. Histology, therefore, continues to play a central role(Alvarez et al. 1999), yet biopsy findings may be inconclusive due to sampling variability or overlapping features with other inflammatory liver diseases. Consequently, a substantial proportion of patients undergo liver biopsy only to receive non-diagnostic or indeterminate results.

Beyond diagnosis, the lack of reliable non-invasive tools to assess ongoing immune-mediated intrahepatic activity further complicates clinical management. Biochemical remission, defined by normal liver enzymes and normal serology, does not always reflect the actual inflammatory process in the liver parenchyma. Thus, many advocate, especially in children, for liver biopsy when reaching remission and stopping the treatment is considered. Remaining inflammation in the liver increased the risk of relapse and poor outcomes(Dhaliwal et al. 2015; Ben Merabet et al. 2021).

The mainstay treatment of AIH is immunosuppression and although it is effective in inducing remission in the majority of patients, long-term management is highly variable and complicated by the lack of reliable non-invasive tools to assess ongoing disease activity. Despite the routine use of biochemical remission, defined by normalization of serum transaminases, IgG levels, and autoantibody titers, as a clinical endpoint, multiple studies have shown that biochemical remission does not consistently reflect histological remission, with up to one-third of patients demonstrating persistent inflammatory activity on biopsy despite normal laboratory values(Laschtowitz et al. 2021)(Feld et al. 2005);(Laschtowitz et al. 2021). This discordance underscores a major limitation in current AIH diagnosis and management: dependence on repeat liver biopsies, which are invasive, subject to sampling error, and not suitable for frequent longitudinal assessment. Furthermore, distinguishing active disease from immune-mediated scarring or quiescent remission solely through serum biomarkers remains a significant unmet clinical need(Cazzaniga et al. 2025; Zhang and Jain 2023). Thus, in most cases, a liver biopsy is needed to confirm histological remission to allow a modification of drug treatment.

Cell-free DNA (cfDNA) liquid biopsies have emerged over the past two decades as a powerful tool for diagnosing and monitoring diseases and enabled their introduction into clinical practice, primarily in cancer(Ignatiadis et al. 2021). Cell-free DNA assays in clinical use are mostly focused on detecting sequence differences (e.g., cancer-specific mutations and transplant-specific genetic variations)(Bianchi and Chiu 2018; Heitzer et al. 2019; Aubert et al. 2024). Recent epigenomic liquid biopsy methods can detect cell type through DNA methylation, which are currently used for early cancer detection (Bruhm et al. 2025). Methods that can probe the transcriptional state of the liver can potentially add critical information on the molecular state of the liver to significantly improve diagnosis accuracy.

We and others have demonstrated that chromatin immunoprecipitation and sequencing of cell-free nucleosomes (cfChIP-seq) can effectively infer transcriptional programs from the cells of origin (Sadeh et al. 2021; Baca et al. 2023). This approach leverages histone 3 lysine 4 tri-methylation (H3K4me3), a well-characterized modification marking transcription start sites (TSS) that is highly predictive of active or poised gene expression (Karlić et al. 2010; Weiner et al. 2015; Liu et al. 2005).

While we initially validated this method in colorectal cancer, liver disease, and cardiology (Sadeh et al., 2021), cfChIP-seq has since been independently adopted across diverse clinical contexts. These include cancer subtyping and bladder cancer profiling (Baca et al. 2023; Lotem et al. 2025), determining gene expression in non-small cell lung cancer (Vad-Nielsen et al. 2020), and monitoring organ-specific stress, such as heart transplant rejection (Jang et al. 2024), exercise-induced tissue damage (Fridlich et al. 2023), and aberrant erythropoiesis in asymptomatic COVID-19 (Ben-Ami et al. 2025).

We previously showed that levels of liver-specific genes detected by H3K4me3 cfChIP-seq increase in correlation with elevated transaminases in liver injury (Sadeh et al. 2021). We thus hypothesized that cfChIP-seq can noninvasively monitor active transcription programs occurring in liver cells of AIH patients, providing additional insights into the disease’s pathogenesis and assisting in the challenging diagnostic process. To test this hypothesis, we serially recruited naive patients referred for diagnostic liver biopsy at two clinics (one focused on pediatric care and the other on adult care). After a complete diagnostic evaluation, ∼30% of the patients with initially elevated transaminase levels had a definite AIH diagnosis (**Fig. 1A-B**).

**Figure 1:**
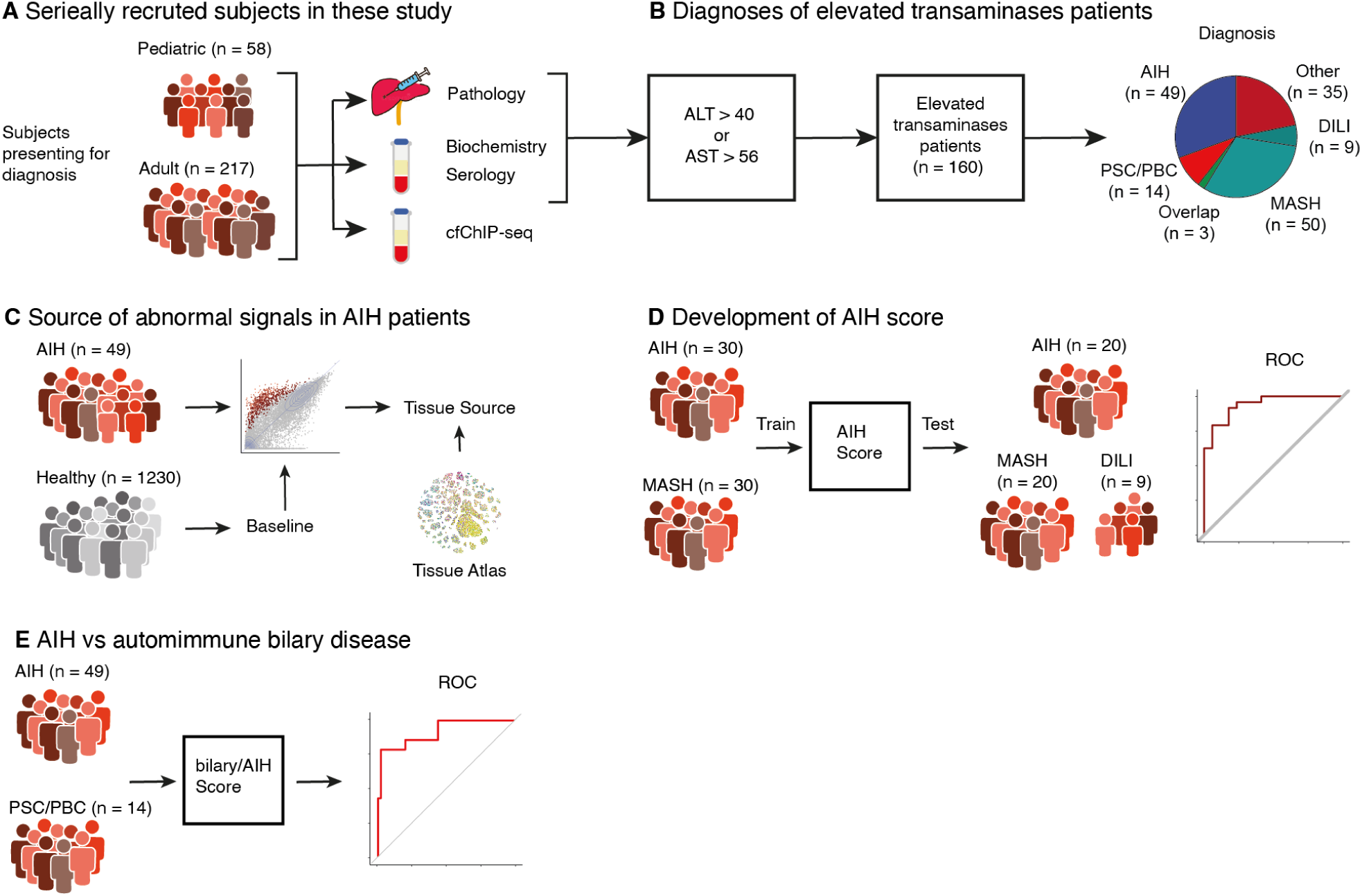
Study outline. A. Plasma samples were collected from patients presenting for diagnostic liver biopsy. In parallel to standard evaluation procedures (serology, biochemistry, biopsy), H3K4me3 cfChIP-seq was performed on 1 ml of plasma. B. Diagnoses of patients with elevated transaminases. C. Comparing each sample against a healthy baseline built from 1230 healthy subjects, we detect genes whose cfChIP-seq signal is significantly above the reference distribution. By contrasting with a tissue atlas of ChIP-seq from the literature, we identify the tissue sources of abnormal signals. D. Using a subset of the patients, we train an AIH score to differentiate the cfChIP-seq signal of AIH patients from MASH patients. We then apply it to classify remaining patients. E. We learn a separate classifier to contrast AIH from autoimmune biliary disease.

We first validated that cfChIP-seq detects abnormal gene expression in AIH patients. By controlling for variable levels of liver damage across the cohort, we identified genes that show elevated signals in AIH patients **(Fig. 1C)**. Using RNA-seq of liver biopsy tissues, we verified that these genes are abnormally expressed in AIH patients’ liver cells compared to other patients’ liver cells. We then trained a cfChIP-seq-based AIH score that combines gene expression levels to differentiate AIH from other liver diseases **(Fig. 1D)**. We demonstrate the score’s utility in a relevant clinical context by evaluating it in a cohort of patients presenting for diagnosis with high transaminase levels. We show that the score distinguishes autoimmune liver diseases from other diseases. Finally, we show that the cfChIP-seq data can distinguish AIH from biliary autoimmune disease **(Fig 1E)**. Together, these results show the potential of this non-invasive method to elucidate the biomarkers and pathogenesis of AIH and to assist in its challenging diagnosis.

## Results

### Elevated hepatocyte-derived cfDNA in AIH plasma samples

We recruited a cohort of patients with potential AIH, defined as elevated transaminases (ALT > 40 and/or AST > 56). We serially recruited patients who underwent diagnostic liver biopsy at two clinics: the Pediatric Gastroenterology unit at Shaare Zedek Medical Center and the adult Liver Clinic at the Hadassah Medical Center (**Table 1**, **Supplementary table 1**). These patients underwent a comprehensive diagnostic assessment, including viral, serological and biochemical testing and needle liver biopsy. AIH was diagnosed based on autoantibodies, elevated IgG, and histological criteria using the IAIH-PG scoring system(Hennes et al. 2008) and excluding other liver diseases (**Table 2**).

**Table 1:** Clinical characteristics of patients presenting for diagnosis with elevated transaminase levels (ALT > 40 U\L and/or AST > 56 U/L).

| Characteristic | AIH<sup>1</sup><br>N = 49 | DILI<sup>1</sup><br>N = 9 | Biliary<sup>1</sup><br>N = 17 | MASH<sup>1</sup><br>N = 50 | Other<sup>1</sup><br>N = 35 | p-value<sup>2</sup> |
| --- | --- | --- | --- | --- | --- | --- |
| Age (years) | 13 (4, 43) | 44 (32, 47) | 38 (34, 50) | 45 (33, 53) | 35 (11, 53) | <0.001 |
| Sex (Female) | 32 (65%) | 4 (44%) | 12 (71%) | 23 (46%) | 17 (49%) | 0.2 |
| ALT (U/L) | 131 (74, 628) | 55 (50, 310) | 107 (68, 170) | 70 (55, 96) | 87 (52, 235) | 0.006 |
| AST (U/L) | 101 (61, 546) | 49 (49, 140) | 82 (48, 130) | 49 (34, 67) | 109 (81, 279) | 0.001 |
| Bilirubin (μmol/L) | 0.8 (0.4, 1.5) | 2.1 (0.8, 7.9) | 1.0 (0.6, 3.5) | 0.9 (0.6, 1.5) | 1.0 (0.5, 2.2) | 0.5 |
| Platelets (10<sup>9</sup>/L) | 279 (214, 331) | 244 (199, 286) | 198 (173, 281) | 244 (203, 280) | 239 (166, 294) | 0.092 |

<sup>1</sup>Median (Q1, Q3); n (%)
<sup>2</sup>Kruskal-Wallis rank sum test; Fisher's exact test

**Table 2:** Further clinical characteristics of AIH diagnosis patients in Table 1.

| Characteristic | AIH (Adults)<sup>1</sup><br>N = 22 | AIH (Pediatrics)<sup>1</sup><br>N = 27 | p-value<sup>2</sup> |
| --- | --- | --- | --- |
| ALT (U/L) | 74 (51, 88) | 475 (169, 1,136) | <0.001 |
| AST (U/L) | 61 (45, 88) | 277 (116, 784) | <0.001 |
| Interface Hepatitis<sup>3</sup> | 14 / 15 (93%) | 20 / 20 (100%) | 0.4 |
| Plasma Cell Infiltration<sup>3</sup> | 11 / 15 (73%) | 24 / 25 (96%) | 0.056 |
| Fibrosis<sup>3</sup> | 14 / 16 (88%) | 13 / 13 (100%) | 0.5 |
| ANA<sup>3</sup> | 9 / 17 (53%) | 12 / 18 (67%) | 0.4 |
| ASMA<sup>3</sup> | 0 / 2 (0%) | 8 / 15 (53%) | 0.5 |
| IGG (g/L) | 1,970 (1,605, 2,066) | 1,180 (982, 1,689) | 0.009 |

<sup>1</sup>Median (Q1, Q3); n / N (%)
<sup>2</sup>Kruskal-Wallis rank sum test; Fisher's exact test
<sup>3</sup>Positive indication

We performed cfChIP-seq with H3K4me3-specific antibody (Sadeh et al. 2021), which enriches cfDNA from active transcription start sites (TSS), on plasma samples from recruited patients (Methods, **Supplementary table 1**). Samples with insufficient yield or specificity were discarded from analysis (Methods; sequencing yield and specificity appear in **Supplementary table S2**). Sequenced reads were mapped to the genome, and the number of normalized reads mapping to each gene’s TSS region was computed, yielding gene counts resembling RNA-seq transcription counts(Sadeh et al. 2021).

Pediatric autoimmune diseases often differ in etiology and clinical presentation from their adult counterparts. Furthermore, physiological baseline profiles can vary by age. We therefore evaluated a cohort of six healthy pediatric individuals against a large cohort of self-identified healthy adults (n = 1,230). The mean signals between the two healthy groups were nearly identical (R = 1), and individual samples exhibited high inter-cohort correlation (median R = 0.975; **Fig. S1A–B**), justifying the use of a shared baseline. Similarly, pediatric and adult AIH patients did not display significant age-related transcriptomic signatures. Principal component analysis (PCA) of genome-wide counts demonstrated comparable profiles between age groups; notably, the primary axis of variation (PC1) was driven by transaminase levels (p = 1.16×10^-11^) rather than age cohort (p = 0.16; **Fig. S1C–D**, ANOVA). Given these comparable profiles, pediatric and adult samples were analyzed as a single unified cohort for all downstream analyses.

Examining plasma samples from AIH patients, we observe many genes with significant levels above the healthy baseline reference (Methods, **Fig. 2A, S2A**). As expected, these genes include many liver-specific genes (**Fig. 2B)**.

**Figure 2:**
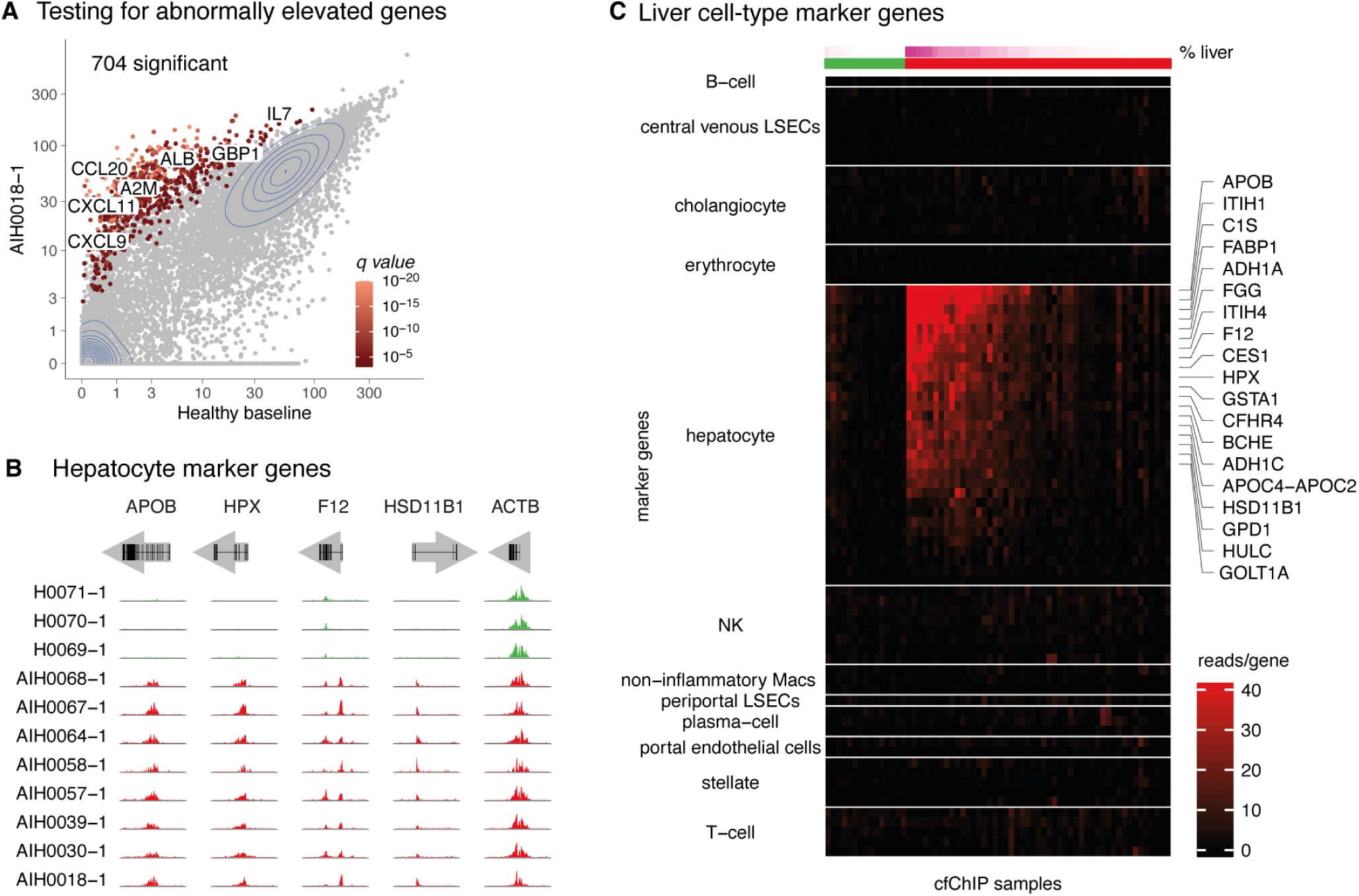
cfChIP-seq displays elevated hepatocyte-derived cfDNA in AIH plasma samples. A. Detection of genes with significantly elevated coverage in a representative AIH plasma sample. For each gene, the mean normalized promoter coverage in the sample (y-axis) was compared with that of a reference healthy cohort (x-axis). Significance indicates whether the observed number of reads in the sample is significantly higher than expected based on the mean and variance in the healthy reference cohort (n = 1230, Methods). B. Genome browser view of cfChIP-seq signal in hepatocytes marker genes (*APOB*, *HPX, F12, HSD11B1*) and *ACTB* as control. Green and red tracks represent healthy and AIH cfChIP-seq samples, respectively. C. Heatmap showing patterns of cfChIP-seq samples (columns) coverage across liver cell-type marker genes (rows). Color represents normalized coverage at the gene promoter. Of all liver cell type markers, a noticeable signal in the AIH samples is observed only in the hepatocyte marker genes.

Linear regression deconvolution (Methods), was consistent with previous studies (Sadeh et al. 2021; Loyfer et al. 2023), where in healthy donors, the major components of cfDNA are hematopoietic cells (neutrophils, megakaryocytes and monocytes), with the liver contributing less than 1% of the cfDNA on average. In contrast, in samples of AIH patients, we see a dramatic increase in liver-derived cfDNAaccounting for a median of 6% and up to 73% of the cfDNA (*t-test p <* 0.000001; **Fig. S2B**). Note that the estimated fractions represent the relative contribution of the tissues to the circulation, not the absolute concentration. Thus, an increase in liver fraction must be compensated by a reduction in the fractions of other tissues, even if their absolute levels remain the same (**Fig. S2B**).

Finally, we examined the set of genes significantly elevated in at least 3 AIH samples, **(supplementary table S3**) against a comprehensive reference atlas comprising 182 H3K4me3 ChIP-seq samples across 36 tissues and cell-types, including solid organs and immune cells (Kundaje et al. 2015; “The International Human Epigenome Consortium: A Blueprint for Scientific Collaboration and Discovery” 2016). These genes showed minimal coverage in the immune cells, the primary source of cfDNA in healthy individuals(Sadeh et al. 2021; Moss et al. 2018), while exhibiting markedly higher enrichment in liver samples (**Fig. S2C**). Enrichment tests of this gene set finds a strong enrichment of the liver (EnrichR human gene atlas *q*<10^-80^; gene overlap 186/618).

To increase the resolution on cfDNA cells of origin obtained from cfChIP-seq in AIH-specific patients, we used gene signatures derived from a single-cell RNA-seq liver atlas ((MacParland et al. 2018), since no such ChIP-seq data exist. Across the ∼10 liver cell-types examined, including hepatocyte, cholangiocyte, endothelial, and immune cells, AIH samples are enriched specifically for hepatocyte marker genes such as *HPX* (hemopexin), *F12* (coagulation factor XII) and *GPD1* (Glycerol-3-Phosphate Dehydrogenase 1) (**Fig. 2C**). The remarkably positive correlation of the hepatocyte marker genes and the liver fraction (*r* = 0.98; **Fig. S2D**) further corroborates the finding that the hepatocytes are the primary source of liver cfDNA in AIH plasma samples.

Comparison of the estimated liver fraction to alanine transferase (ALT) levels measured in time-matched blood samples, displays a good agreement between the two modalities despite the differences in half-life of these analytes(Giannini et al. 2005; Lo et al. 1999) (*r* = 0.85; *p* < 6×10^-11^; **Fig. S2E**). When performing principal component analysis (PCA) of the samples (Methods) the first principal component (which accounts for 25% of variability) is highly correlated with estimated liver fraction (*r =* 0.92, *p* < 2×10^-16^. **Fig. S2F**).

These findings indicate that the predominant abnormality in AIH circulating DNA is an increase in hepatocyte-derived DNA. The autoimmune nature of the disease suggests that this increase is due to an immune attack on hepatocytes.

### cfChIP-seq identifies hepatocyte response in AIH patients

AIH is characterized by a complex process that involves the activation of CD4+ effector and regulatory T cells, cytokine and chemokine production, and more(Webb et al. 2018). To explore whether cfChIP-seq can detect these processes in plasma, we leveraged the fact that H3K4me3 marks transcription start sites and is closely linked to RNA polymerase activity ((Li et al. 2007);(Karlić et al. 2010; Wang et al. 2023)). Consequently, cell-free H3K4me3 levels reflect the transcriptional state and the relative number of active cells contributing to the circulation.

In our cohort, changes in these levels are driven by two major processes. First, a change in the cell-type composition of the cell-free DNA pool changes the number of cells where a gene is marked, thus changing its level (e.g., ALB levels rise with increased hepatocyte cells contribution). Second, a transcriptional change within one (or more) cell types can lead to change in the promoter marks (e.g., response to viral infection leads to increased activity of interferon response genes in affected cells).

To distinguish changes within specific cell types against a background of changes in cell-type composition (Shen-Orr and Gaujoux 2013), we devised the following strategy: First, we apply deconvolution to estimate each sample’s cell-type composition (Methods). For each sample, we construct a composition-informed reference that accounts for the relative composition and the estimated mean and variance of gene levels in each cell type. By comparing this sample-specific reference with the observed values in the sample, we identify genes that are significantly above or below the composition-informed reference (**Fig. 3A**; Methods).

**Figure 3:**
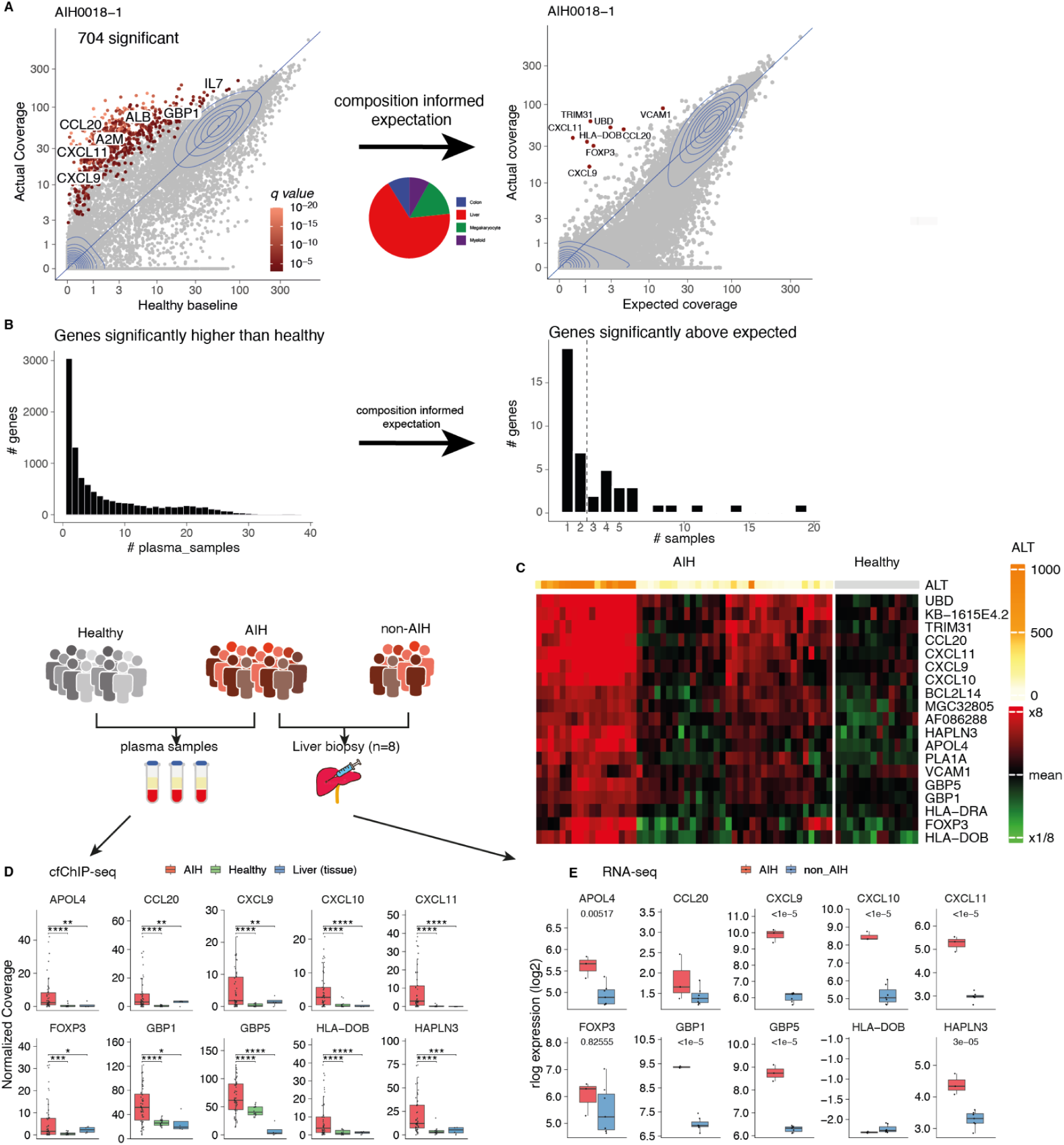
cfChIP-seq deconvolution reveals an active immune transcription program in AIH hepatocytes. A. Composition-informed normalization of cfChIP-seq signals. Scatter plots of a representative AIH sample (AIH0018) comparing observed gene coverage against a healthy baseline (left; same as Fig. 1B) and against a sample-specific expected reference (right). The expected reference is constructed via linear regression deconvolution to account for individual cell-type composition. Red points indicate genes significantly elevated (q < 0.01) above the composition-informed mean and variance, identifying active transcriptional changes beyond shifts in cell-of-origin. B. Global reduction of elevated signals following composition correction. Distribution of significantly elevated genes across the AIH cohort before (left; same as Fig. S1E) and after (right) composition-informed correction. Histograms show the number of genes (y-axis) as a function of the number of samples in which they are elevated (x-axis). Correction for tissue composition isolates a specific subset of genes reflecting active transcriptional programs. C. AIH-specific hepatocyte inflammatory signature. Heatmap of genes exhibiting significantly elevated H3K4me3 levels after composition correction in at least two AIH samples. Rows represent genes and columns represent individual plasma samples. Color scale indicates gene coverage compared to the mean coverage of that gene across samples. Note the consistent induction of pro-inflammatory and interferon-responsive genes (e.g., *CXCL9-11*, *GBP1/5*, *UBD*) across the cohort. D. Distribution of normalized promoter coverage shown in panel C. The median and distribution of normalized coverage in cfChIP-seq for AIH and healthy samples, and ChIP-seq of liver tissue from reference atlas, are shown. E. Median and distribution of RNA-seq rlog-transformed expression of genes shown in panel C data from liver biopsies of patients with AIH and individuals without AIH.

Applying this strategy to AIH samples, shows that the vast majority of the 896 genes with significantly elevated signals in multiple AIH samples (**Fig. 1D)** do not deviate substantially from composition-informed reference, suggesting that they are consistent with stereotypical liver chromatin patterns (**Fig. 3B**). However, a close inspection reveals a set of 20 genes with levels significantly above expected in several samples (**Fig. 3C, S3A**). This group includes the *CXCL9-11* (C-X-C motif chemokine ligand) genes, which are expressed in inflamed hepatocytes and play a role in the AIH immune response and liver fibrosis(Czaja 2014; Ikeda et al. 2014; Christen and Hintermann 2016; Zeremski et al. 2011). Additional genes in this group are *UBD* (Ubiquitin D) and *TRIM31* (Tripartite Motif Containing 31), which are induced by pro-inflammatory cytokines, and *HLA-DOB* (Major histocompatibility complex, class ll, Do Beta), which was identified as affecting the occurrence and development of hepatitis B (HBV) but not reported in the context of AIH to the best of our knowledge(Raasi et al. 1999; Li et al. 2022; Jia et al. 2021). The interferon-responsive genes *GBP1* and *GBP5,* whose expression is associated with liver injury and inflammation(Xiao and Tipoe 2016; Ding et al. 2022; Milks et al. 2009), were also significantly above expectation. Another gene identified is *HULC* (highly upregulated liver cancer), which is expressed in normal hepatocytes but is strongly induced in hepatocellular carcinoma and HBV infection, and is involved in inflammatory injury in rats with cirrhosis(Matouk et al. 2009; Panzitt et al. 2007; Zhu et al. 2019). Notably, many of these genes, which have high coverage in AIH samples, carry no H3K4me3 signal in normal liver nor any of the other tissues represented in the reference atlas, indicating that they reflect activation of an abnormal transcription program in patients with AIH (**Fig. 3D; S3B**).

To rule out the possibility that these genes reflect a transcriptional program in cell types other than the liver, we tested the correlation between the gene levels and the estimated tissue fraction across all tissues composing the samples. This analysis revealed that these genes are positively correlated to the estimated liver fraction in the AIH samples and negatively correlated to all other tissues (**Fig. S3C-D**). The results suggest that the activity levels of these genes are strictly coupled to hepatocyte death, most likely reflecting transcription in the hepatocytes of patients with AIH.

To further validate the activity of these genes in AIH liver, we performed RNA-seq on liver biopsies (Methods). The patients were referred to liver biopsy as part of their clinical treatment. The pathology report and subsequent clinical trajectory identified clear cases of AIH (n = 3), metabolically associated steatohepatitis (MASH) (n=2), nonspecific findings (n=3) and normal biopsy (n=2). The RNA-seq data were processed and normalized (Methods). Examining the levels of the AIH-induced genes described above (Fig. 3E), we see that indeed 9/20 are significantly over-expressed in RNA-seq from AIH vs. Non-AIH liver biopsy (q < 0.05, **Supplementary table 6**, Methods).

These results strongly suggest that the AIH-induced genes identified above reflect an immune-mediated hepatocyte death within the livers of AIH patients.

### Plasma-based AIH score

We next examined the clinical relevance of the cfChIP-seq signals discussed above. In a clinical setting, a patient presenting with elevated liver transaminases often requires a liver biopsy, primarily to exclude AIH. At the same time, other diagnoses such as MASH, drug-induced liver injury (DILI), PBC (Primary biliary cholangitis), Wilson’s disease, and various forms of viral hepatitis can be diagnosed with other means(Russo 2020; Mack et al. 2020; Czaja et al. 2002). Because AIH has a specific treatment, distinguishing it from other etiologies with elevated liver enzymes is clinically important.

We expanded our analysis to the full set of plasma samples from the cohort described above and added samples of other clinical situations with increased liver damage (**Supplementary Table 1**). The cohort comprises patients with MASH (n = 89), DILI (n = 14), cholestatic liver disease (PSC/PBC, n = 21), patients recovering from liver transplantation (LTx, n = 7), overlap syndrome (AIH and PBC or PSC, n = 8) and other diseases (n = 43).

We focus on samples with elevated aminotransaminase levels (ALT> 40 U/L or AST > 56 U/L) in whom AIH was included in the differential diagnosis. The standard diagnosis criteria are to perform biochemical (alkaline phosphatase, GGT, and bilirubin), serological tests (Antimitochondrial antibodies (AMA), Antinuclear Antibody (ANA), Anti-Smooth Muscle Antibody (ASMA), Liver/Kidney Microsomal antibody (LKM), and Anti-Soluble Liver Antigen (anti-SLA), immunoglobulins (IgG and IgM), and viral testing (HBV, HCV) (European Association for the Study of the Liver 2025).

We reasoned that to distinguish AIH from other liver diseases, we can train a classifier between the AIH cohort and a coherent cohort of non-AIH, ideally of one disease with a different etiology. We decided to use the MASH cohort that had a large number of patients (50 patients with elevated transaminases), involved inflammation, but due to a different etiology, and exhibited increased levels of serum transaminases. We randomly split both the AIH and MASH samples into training and validation groups (n = 60% and 40%, respectively; **Supplementary Table 1**).

We performed our analysis on genomic regions with residual signals above the expected levels according to tissue composition (as in **Fig 3A**). However, we expanded the analysis to include all H3K4me3 peaks, rather than just the promoters of annotated genes (Methods). We used leave-one-out cross-validation on the training set to identify genomic regions with a significant increase in the residual signal of AIH compared to MASH samples (*t*-test, *q* < 0.1) and to optimize the number of regions to use. This optimization resulted in choosing 30 regions (Methods). Many of these regions lack signal in almost all MASH samples (**Fig. 4A**), supporting the role of these genes in an AIH-unique immune response. We define the AIH score as the sum of signals across these regions in each sample (**Supplementary table 7**).

**Figure 4:**
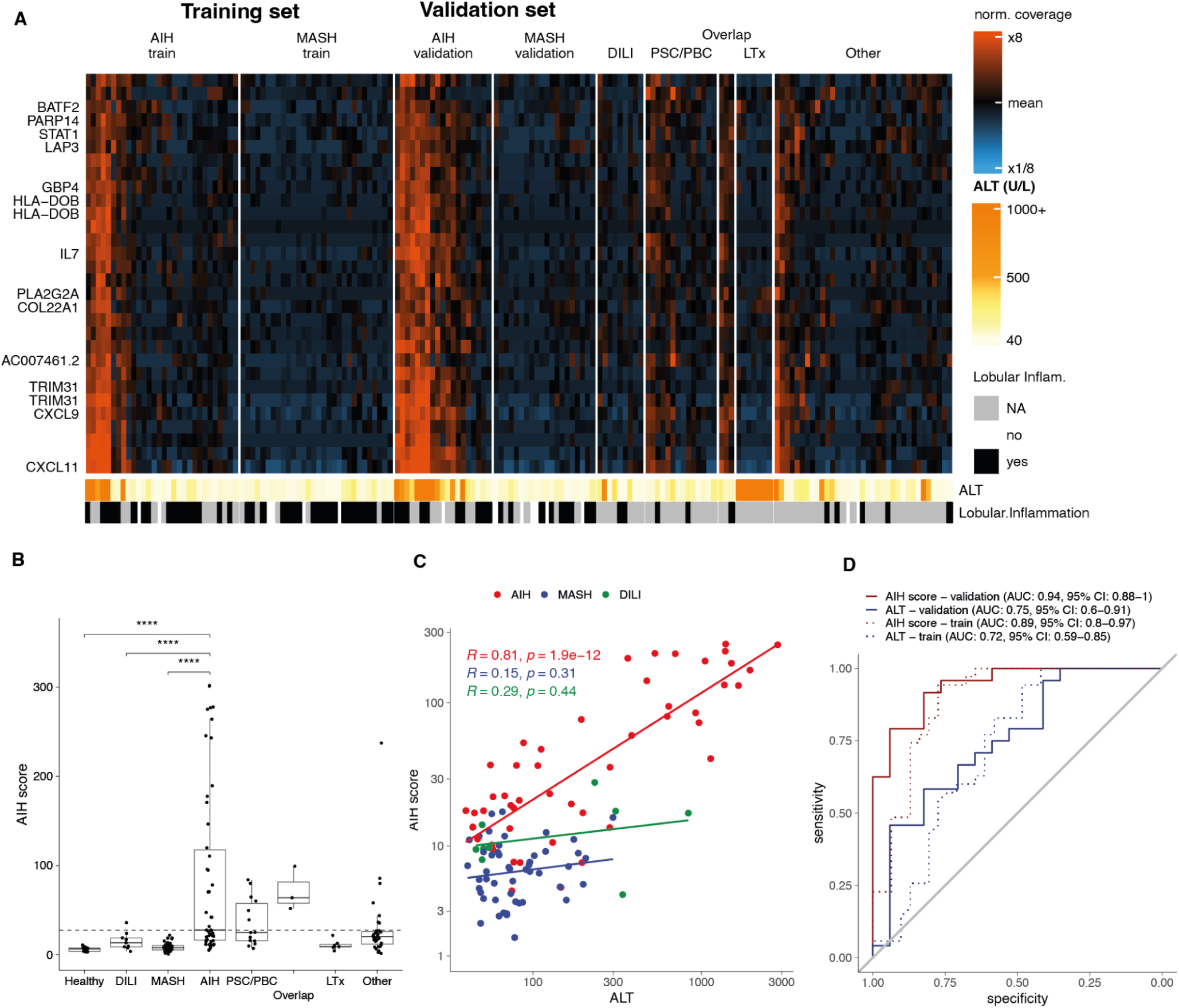
plasma-based classifier for AIH diagnosis. A. Heatmap showing patterns of the cfChIP-seq coverage in genomic regions that make up the AIH score across different liver conditions. Color scale represents fold change to the mean of all samples. Attribute bars below heatmap show serum alanine aminotransferase (ALT) level at the date of sample collection and presence of lobular inflammation in histology (when available). B. Median and distribution of the AIH-score across multiple cohorts. The score (y-axis) indicates the difference in the cumulative coverage of the specific regions shown in A. C. AIH score increases with serum alanine aminotransferase (ALT) levels in AIH patients but not in MASH and DILI. AIH score (y-axis) as a function of ALT (x-axis). cfChIP-seq samples (dots) are colored by group lines indicating the linear fit of each group. D. Classifier performance for contrasting AIH vs MASH+DILI among samples with elevated transaminase levels. ROC of AIH score and ALT performance on training and validation cohorts.

The AIH score significantly distinguishes AIH samples from MASH samples as well as healthy subjects and DILI patient samples, which were not part of the training process (**Fig. 4B**).

Most AIH-related signals are associated with hepatocyte cell death. Thus, we compare the computed score to the ALT levels of the corresponding samples. Within the AIH samples the two measures have a linear relationship, consistent with the liver-death coupled nature of the AIH signals (and the residuals). In contrast, this phenomenon is much less pronounced in MASH and DILI samples, suggesting that this signature captures a transcription program typically inactive in the non-AIH liver (**Fig. 4C**).

We evaluated the AIH score-based classifier’s ability to discriminate between AIH and MASH/DILI plasma samples on the training set (area under the curve (AUC) = 0.89) and the validation set (AUC = 0.94; **Fig. 4D, S4A**). This discrimination outperforms ALT as an indicator (AUC = 0.72, and 0.75, on training and validation cohorts, respectively; **Fig. 4D, S4B**).

### AIH score identifies autoimmune liver injury

To better understand the physiological relevance of AIH score, we examined AIH scores across the diverse cohort of liver diseases (**Fig. 4B**). The low signals observed in samples of patients with MASH, including those with marked inflammation (**Fig. 4A, S5**), suggest that liver injury alone is insufficient to induce this signature. We observe intermediate AIH scores in patients with PBC and PSC, autoimmune biliary duct diseases (**Fig. 4A-B**). To further define the specificity of this signature, we performed a post-hoc analysis of patients in the heterogeneous ‘Other’ disease category who exhibited AIH scores above the median of the AIH cohort (**Fig. 4B**). This subset was characterized by intense immune or inflammatory activity; notably, the highest signal was observed in a pediatric patient with acute severe hepatitis and imminent liver failure which eventually was diagnosed with Cytomegalovirus virus (CMV) hepatitis. CMV infection is known to modulate immune responses and has been implicated in the pathogenesis of various autoimmune diseases via molecular mimicry, inflammation, and nonspecific B-cell activation (Gugliesi et al. 2021; Halenius and Hengel 2014). Other high-scoring samples included patients with Hepatitis B virus (HBV) (concomitant with elevated IgG and anti-smooth muscle antibodies), undefined metabolic disease, and juvenile rheumatoid arthritis and elevated liver enzymes with non-specific findings in liver biopsy. One outlier exhibited a complex serological profile positive for IgG4, ASCA, anti-mitochondrial antibodies, and anti-gluten antibodies. Beyond the above patients, only one high-scoring patient lacked any documented immune-related diagnosis.

Together, these results suggest that the elevated AIH scores reflect an autoimmune-mediated hepatocyte death, rather than nonspecific inflammation or hepatocyte injury.

### AIH score complements clinical diagnostic measures

We next assessed the behavior of the AIH score across clinically relevant subgroups of autoimmune liver disease. Seropositive and seronegative AIH patients exhibited comparable score distributions across ALT levels (**Fig. S6A–B**), suggesting that the cfChIP-seq–derived signal is not dependent on circulating autoantibodies. There is partial concordance between the AIH and International Autoimmune Hepatitis Group (IAIHG) scores; however, several patients with low or indeterminate IAIHG scores showed elevated AIH scores (**Fig. S6C**), suggesting that cfChIP-seq may capture aspects of immune-mediated hepatocellular injury not fully reflected by conventional scoring systems. Notably, comparison of the AIH score with serum IgG levels revealed no significant correlation (**Fig. S6D**).

We next examined the relationship between the AIH score and histopathological features commonly used to define disease activity. Samples with histological evidence of interface hepatitis or prominent plasma cell infiltration tended to have higher AIH scores than samples lacking these features (**Fig. S6E**). However, the number of cases without interface activity or plasma cell infiltrates was limited, restricting the statistical power of these comparisons.

These findings indicate that the AIH score provides complementary information to existing clinical, serological, and histological assessments. By directly capturing liver-specific autoimmune activation from plasma, cfChIP-seq provides a valuable adjunct for evaluating suspected liver autoimmune diseases, particularly in diagnostically ambiguous cases or when biopsy interpretation is inconclusive.

### Cell-type-based classification of autoimmune liver diseases

While the AIH score effectively distinguishes AIH from non-autoimmune liver diseases, it does not inherently differentiate between primary hepatocyte damage (characteristic of AIH) and autoimmune-mediated biliary duct injury (cholangiocyte damage). In clinical practice, distinguishing AIH from Primary Sclerosing Cholangitis (PSC) or Primary Biliary Cholangitis (PBC) remains a diagnostic challenge, often requiring a combination of invasive liver biopsies, magnetic resonance cholangiopancreatography (MRCP), and specific serological markers like alkaline phosphatase (ALP) and anti-mitochondrial antibodies (AMA). Even with these tools, “overlap syndromes”—where patients exhibit features of both hepatitis and cholestasis—frequently complicate definitive diagnosis and treatment selection.

We reasoned that cfChIP-seq could provide a more granular resolution of the specific cell types under immune attack by identifying gene signatures unique to each lineage. By comparing the cfChIP-seq profiles of AIH patients against those with PSC and PBC, we identified two distinct gene sets: a “AIH-specific” signature, preferentially enriched in AIH, and a “PSC/PBC-specific” signature, enriched in biliary disease samples (**Fig. 5A**; Methods). The cumulative signal across these sets was used as a score. Interestingly, *KR7*, one of the biliary-specific genes identified, is a hallmark for cholangiocyte transcription(Aizarani et al. 2019).

**Figure 5.**
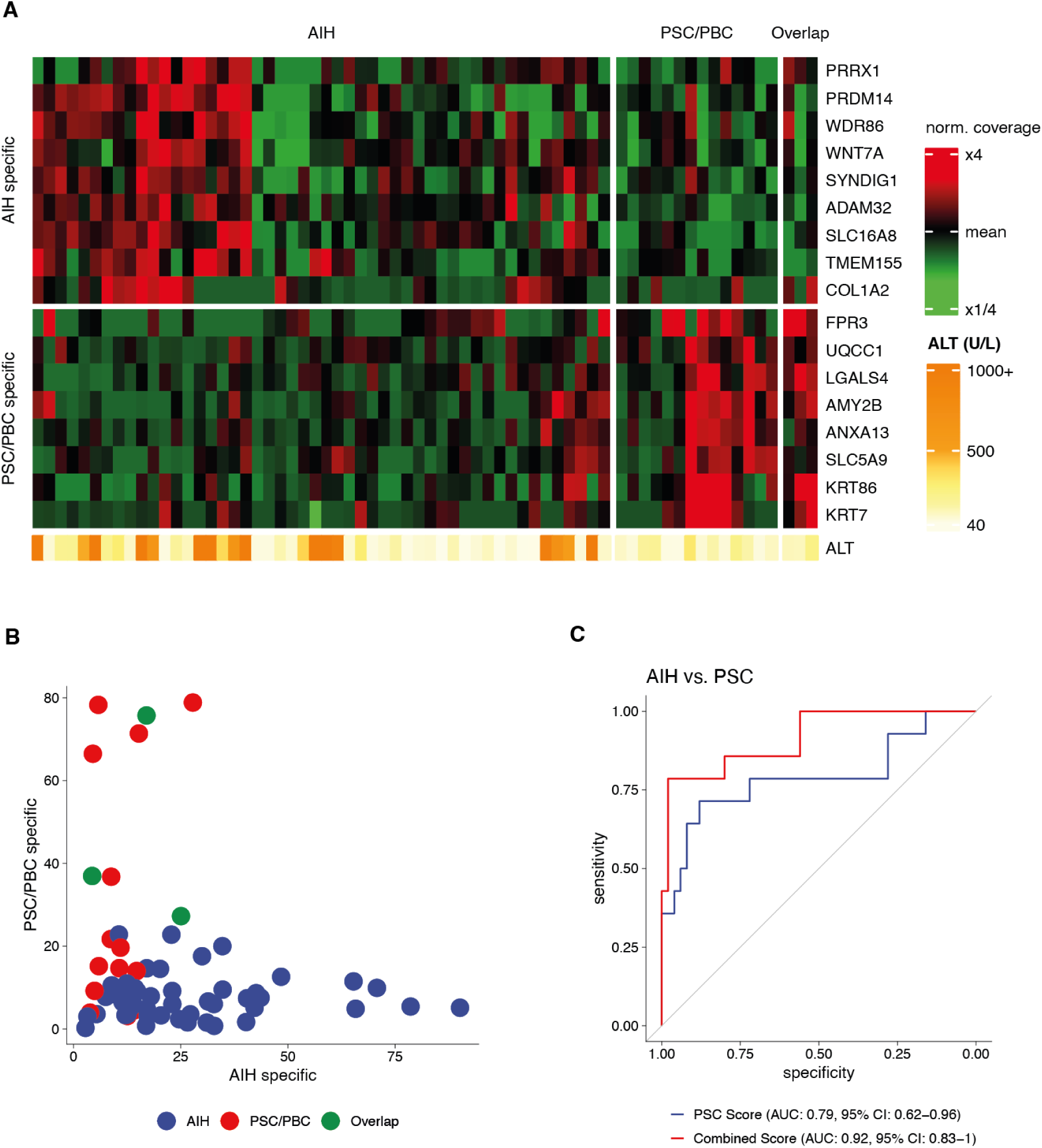
Differential classification of autoimmune liver diseases based on cell-type-specific signatures. A. Heatmap of AIH-specific and PSC/PBC-specific gene enrichment across plasma samples of relevant groups. B. Scatter plot of “AIH specific” versus “PSC/PBC specific” by disease group. AIH and biliary samples show distinct polarization, with overlap cases displaying intermediate signals. C. ROC curves for the PSC/PBC signature and a combined model that includes also the AIH signature (AUC = 0.92) for the discrimination of AIH from biliary disease.

These metrics effectively separated AIH from PSC/PBC (**Fig. 5B**). Specifically, AIH samples were characterized by high AIH-specific and low biliary-specific scores, whereas biliary samples showed the inverse. A composite classifier utilizing both signatures achieved an AUC of 0.92 for group discrimination (95% CI 0.83-1; **Fig. 5C**). Given the limited sample size of PSC/PBC and overlap cases in our current cohort, these results provide a promising proof-of-concept that cell-type-specific signals in cfChIP-seq can facilitate the non-invasive differential diagnosis of autoimmune liver diseases.

### AIH score dynamics in treated patients and biochemical remission

To examine the behavior of our score during AIH treatment, we also included samples from AIH patients at biochemical remission, either under immunosuppressive therapy or after weaning. Across the cohort, treated AIH samples exhibited significantly lower AIH scores compared with samples of patients not receiving active immunosuppression, consistent with effective immunosuppressive control of immune-mediated hepatocyte injury at the group level (**Fig. 6A**). This reduction was not attributable to the reduction in transaminase levels alone (**Fig. 6B**), but instead reflected attenuation of the AIH-associated transcriptional signature captured by cfChIP-seq.

**Figure 6:**
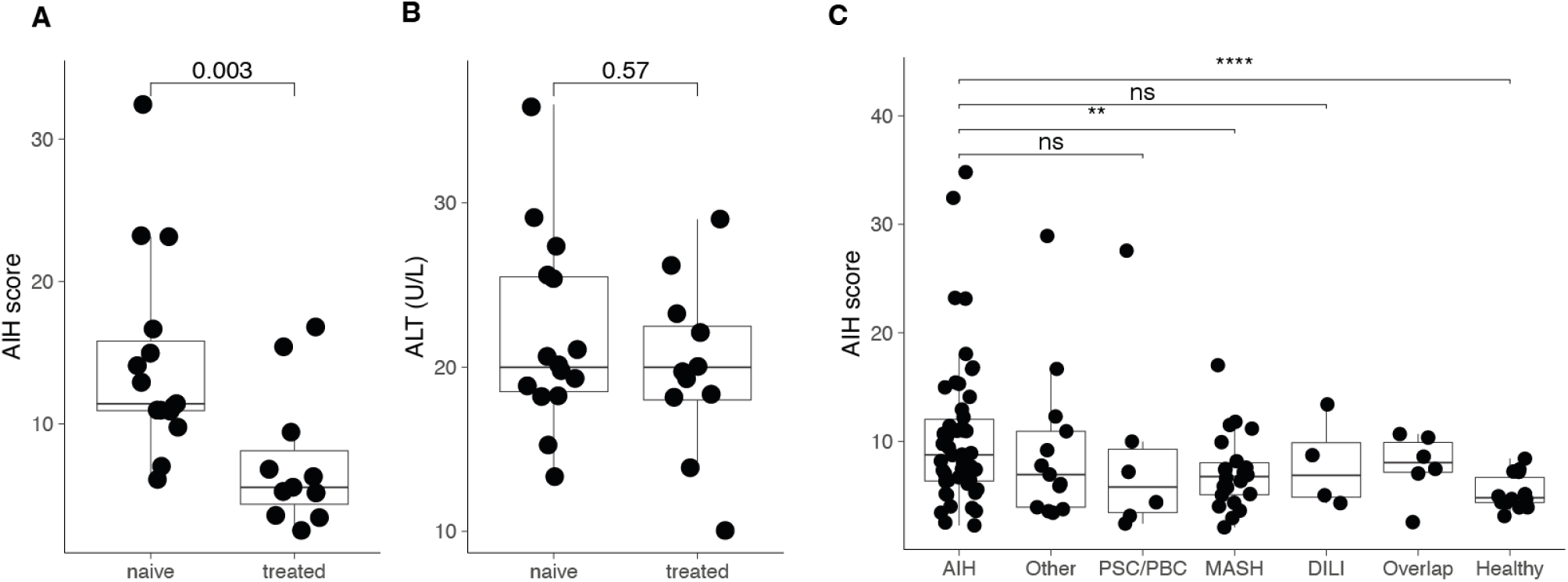
AIH score dynamics in patients at biochemical remission. A. Median and distribution of the AIH-score in treatment-naïve and treated AIH samples. At the group level. B. Same as A for serum alanine aminotransferase. Treated patients’ samples exhibit a significant reduction in AIH score compared with samples from patients not under treatment, accompanied by normalization of ALT values, consistent with treatment response. C. Median and distribution of the AIH-score across disease groups. Elevated AIH scores are observed specifically in AIH samples, including a subset of samples obtained during biochemical remission, whereas scores remain low in other liver diseases and in healthy controls.

We next examined the behavior of the AIH score in samples obtained during biochemical remission, defined as normalization of serum ALT and AST levels. As control, we included samples from patients with other conditions that had normal transaminase levels. In the majority of remission samples, AIH scores decreased to levels comparable to those observed in non-AIH inflammatory liver diseases and healthy plasma, indicating resolution of the immune-associated hepatocyte signal. However, a subset of remission samples retained elevated AIH scores despite normal transaminase levels. Notably, this pattern was specific to AIH. It was not observed in samples from patients with DILI, MASH, or healthy controls, all of which showed uniformly low AIH scores under comparable biochemical conditions (**Fig. 6C**).

This dissociation between biochemical markers and cfChIP-derived AIH scores suggests that cfChIP-seq captures immune-mediated transcriptional activity that may persist despite apparent biochemical quiescence. This difference is clinically relevant, as biochemical remission is an imperfect indicator of clinical remission. In one patient with normal transaminase levels, a persistently elevated cfChIP-seq AIH score indicated ongoing immune activity, which was subsequently corroborated by liver histology demonstrating active disease.

Together, these findings indicate that while AIH scores generally normalize with effective treatment and biochemical remission, a subset of patients may exhibit persistent elevations that may reflect ongoing immune-mediated hepatocyte activity not captured by standard laboratory tests. This phenomenon appears specific to autoimmune liver disease and supports the potential utility of cfChIP-seq as a complementary tool for disease assessment beyond conventional biochemical markers.

## Discussion

Accurate, non-invasive diagnosis of liver disease etiology remains a major unmet need in hepatology. Current diagnostic tools—biochemical markers, serology, imaging, and scoring systems—largely reflect downstream consequences of liver injury and often fail to capture the molecular processes occurring within affected liver cells. As a result, liver biopsy remains central to the diagnosis and classification of many liver diseases, particularly autoimmune conditions, despite its invasiveness, sampling variability, and limited suitability for longitudinal monitoring. In this study, we demonstrate that plasma cfChIP-seq provides a different diagnostic modality by directly reporting cell-type–specific transcriptional activity in dying liver cells and that this information can be used to distinguish autoimmune liver diseases from other causes of liver injury.

Autoimmune hepatitis (AIH) represents an especially challenging diagnostic context in which to evaluate such a technology. AIH is a chronic liver disease with severe outcomes. It is defined not merely by hepatocyte death but by immune-mediated activation of hepatocytes and intrahepatic immune signaling, features that are incompletely captured by standard serum biomarkers. Despite advances in elucidating the underlying immunopathogenic mechanisms, diagnosis remains challenging and traditionally requires an invasive liver biopsy for histological confirmation(Manns et al. 2010; “International Autoimmune Hepatitis Group Report: Review of Criteria for Diagnosis of Autoimmune Hepatitis” 1999). Liquid biopsy is an emerging field of medical diagnostic technology, and considerable effort has been made over the past two decades to develop assays that can replace tissue biopsies with non-invasive liquid biopsy alternatives. However, most breakthroughs in the past few years have occurred in fields where DNA sequence changes are available, such as oncology and prenatal genetic screening(Ignatiadis et al. 2021; Alix-Panabières and Pantel 2021). More recent advances have demonstrated the capacity of cfDNA to reflect abnormal cell death in somatic tissues (Moss et al. 2018; Sadeh et al. 2021). However, little is known about how liquid biopsies capture transcriptional programs that reflect the functional state of the cells of origin. This is notably missing in the context of autoimmune hepatitis (AIH), which is distinguished from other liver diseases not in the type of dying cells but rather in their state. Using AIH as a biological and clinical model, we show that cfChIP-seq detects both the magnitude of hepatocyte injury and, critically, the immune transcriptional programs that distinguish autoimmune liver disease from metabolically driven, toxic, or nonspecific inflammatory injury.

Beyond binary classification of autoimmune versus non-autoimmune liver disease, cfChIP-seq provided additional resolution within autoimmune liver disorders themselves. By exploiting lineage-specific transcriptional signatures, we were able to distinguish autoimmune injury characteristic of AIH from cholangiocyte-directed injury seen in PBC and PSC. This cell-type–resolved information is difficult to obtain noninvasively and underscores the unique value of cfChIP-seq for phenotyping complex autoimmune liver disease.

We find that cfChIP-seq identifies increased hepatocyte cell death in patients with active disease, which correlates with elevated serum transaminase levels. We devised a statistical model to account for this major confounder and successfully isolated the abnormal gene activation in dying cells. The abnormally activated genes include members of the C-X-C motif chemokine ligand family (CXCL9/10/11), which have been implicated in AIH responses in hepatocytes and in liver fibrosis (Zeremski et al. 2011; Czaja 2014). Other genes (e.g., guanylate-binding proteins 1/5, GBP1/5) are known to play inflammatory roles in various liver diseases(Xiao and Tipoe 2016; Ding et al. 2022; Milks et al. 2009) but have not been reported in AIH. Additional genes (e.g., *TRIM31, UBD)* showed elevated levels in the AIH cohort and also in cfChIP-seq samples of patients with other liver diseases. These genes, which are involved in the ubiquitin-proteasome pathway (Song et al. 2016), presumably reflect a hepatocyte stress response. Importantly, RNA-seq of matched liver biopsies confirmed that a substantial subset of these genes is transcriptionally upregulated in AIH liver tissue, providing direct biological validation that cfChIP-seq captures intrahepatic immune activity.

Beyond studying underlying processes in AIH, we also sought to leverage this assay to assist in AIH diagnosis. This was achieved by identifying AIH-specific cfChIP signals that were absent in the control group of samples from patients with other liver diseases (primarily MASH patients). We show that this classifier is effective in distinguishing AIH and related diseases (overlap syndrome) from DILI, MASH, and other diseases.

From a clinical perspective, cfChIP-seq has the potential to address several unmet needs in the evaluation of suspected autoimmune liver disease. In patients presenting with elevated transaminases, a plasma-based cfChIP-seq assay could provide early, non-invasive evidence of autoimmune-mediated liver injury and help prioritize patients for prompt liver biopsy and immunosuppressive therapy. This is especially true in patients with negative autoimmune serology or an equivocal liver biopsy.

For example, during the study period, several cases were identified in which retrospective application of our classifier could have influenced clinical management. For instance, a 3-year-old girl presented with acute hepatitis and impending liver failure and underwent liver biopsy. Empirical steroid therapy initiated immediately after biopsy resulted in improved liver enzyme levels; however, steroid treatment was discontinued after histology was deemed inconsistent with autoimmune hepatitis (AIH). Several months later, liver enzymes increased again, and a repeat biopsy confirmed AIH. Retrospective analysis using the cfChIP-seq classifier from the initial presentation was unequivocally compatible with AIH. This case highlights the clinical value of supplementary diagnostic tools, even when a liver biopsy has been performed, in facilitating accurate and timely diagnosis.

The clinical relevance of cfChIP-seq is further illustrated by its behavior in treated AIH patients. In most cases, AIH-specific cfChIP signals declined with immunosuppressive therapy, paralleling biochemical improvement. However, a subset of patients in biochemical remission retained elevated AIH scores, a pattern not observed in other liver diseases. This dissociation mirrors the known discordance between biochemical and histological remission in AIH (Mack et al. 2020) and suggests that cfChIP-seq may detect ongoing immune-mediated hepatocyte activity that escapes conventional laboratory assessment. Such capability could be particularly valuable for guiding treatment tapering and identifying patients at risk for relapse(Dhaliwal et al. 2015; Ben Merabet et al. 2021).

Several limitations should be acknowledged. First, while cfChIP-seq provides a powerful window into disease-associated transcriptional programs, its implementation in routine clinical practice remains nontrivial. This is similar to other current epigenomic liquid biopsies. There is a clear path to implementation in the clinic, involving commercialization and standardization. The aim here is to establish the potential utility that justifies these efforts. Moreover, the present study was not designed to assess the utility of cfChIP-seq for longitudinal disease monitoring, treatment response assessment, or guidance of immunosuppressive dose adjustments. Although we observed suggestive signals in some patients in biochemical remission, systematic evaluation of cfChIP-seq dynamics over time, including during treatment initiation, tapering, or flare, remains beyond the scope of this work. Addressing these aspects will require larger, prospective studies and represents an important direction for future research.

In conclusion, this study positions cfChIP-seq as a liquid biopsy platform that extends beyond the detection of liver injury to capture disease-defining transcriptional programs within hepatocytes. Using autoimmune hepatitis as a clinically and biologically rigorous test case, we demonstrate that cfChIP-seq enables accurate, non-invasive discrimination of autoimmune liver diseases and provides insights into cell-type–specific immune activity that are difficult to obtain by existing methods. These findings support the development of cfChIP-seq as a molecular “liquid liver biopsy” with potential applications in diagnosis, disease stratification, and treatment monitoring in autoimmune liver disease.

## Methods

### Patients

Plasma samples of patients and healthy controls were collected in the pediatric Gastroenterology Institute at Shaare Zedek Medical Center (SZMC) and at the Hadassah Medical Organization - Hebrew University Hospital (HMO). The samples were taken from patients under various clinical conditions: (1) patients undergoing liver biopsy for AIH diagnosis or other liver disease due to persistent elevation of liver enzymes or (2) to establish histological remission in patients with established AIH under treatment or (3) patients with established AIH under treatment with no adjacent liver biopsy. The control group comprises patients with normal liver biopsy or with no elevation of liver enzymes nor other liver disease. The study was approved by the Ethics Committees of the SZMC (0269-19-SZMC) and HMO (0752-21-HMO). Informed consent was obtained from all individuals or their legal guardians before blood sampling. The sample size was determined by the availability of samples.

### Patient and public involvement

All patients were offered to receive the results of the study but were not involved in the development or the design of the study nor in the recruitment or conduction of the study.

### Plasma cfChIP-seq

#### Immunoprecipitation, NGS library preparation, and sequencing

Sample collection and handling, Immunoprecipitation, library preparation, and sequencing were performed by Senseera LTD. as previously reported(Sadeh et al. 2021), with certain modifications that increase capture and signal-to-background ratio. Briefly, ChIP antibodies were covalently immobilized to paramagnetic beads and incubated with plasma. Barcoded sequencing adaptors were ligated to chromatin fragments and DNA was isolated and next-generation sequenced.

#### Sequencing Analysis

Reads were aligned to the human genome (hg19) using bowtie2 with ‘no-mixed’ and ‘no-discordant’ flags. We discarded fragment reads with low alignment scores (-q 2) and duplicate fragments. See **Supplementary Table 2** for read number, alignment statistics, and numbers of unique fragments for each sample.

Preprocessing of sequencing data was performed as previously described. Briefly, the human genome was segmented into windows representing TSS, flanking to TSS, and background (rest of the windows). The fragments covering each of these regions were quantified and used for further analysis. Non-specific fragments were estimated per sample and extracted resulting in the specific signal in every window. Counts were normalized and scaled to 1 million reads in healthy reference accounting for sequencing depth differences. Detailed information regarding these steps can be found in the supplementary note (Sadeh et al. 2021). Samples with fewer than 400,000 unique reads or less than 25% signal were discarded from analysis.

### Tissue RNA-seq

#### RNA extraction, NGS library preparation, and sequencing

##### Liver Tissue Processing and RNA extraction

Formalin-fixed paraffin-embedded (FFPE) liver sections of 10-20μm thickness were deparaffinized in xylene followed by ethanol rehydration. The deparaffinized tissues were then air-dried for 5 min and transferred to pre-chilled 1.5 ml tubes.

Samples were then incubated in 150μl FFPE-specific lysis buffer (PKD buffer, QIAGEN) with proteinase K (20 mg/ml) at 56°C for 15 min, followed by incubation at 80°C for 15 min to reverse formaldehyde crosslinks. Lysates were centrifuged (14,000 × g, 15 min, RT) and supernatants were collected. Genomic DNA was removed via DNase digestion (DNase I, 5 U per sample; 15 min, RT).

##### RNA purification

Nucleic acids were isolated using silica-membrane columns (RNeasy MinElute® Spin Columns, QIAGEN). Lysates were mixed with 100% ethanol, loaded onto columns, and washed according to manufacturer’s protocol. RNA was eluted in 30μl RNase-free water and quantified using a Qubit 4.0 Fluorometer (ThermoFisher Scientific). Purity (A260/A280:1.8–2.0) and integrity (DV ≥ 50%) were assessed via TapeStation 4200 (Agilent Technologies). Samples with ribosomal integrity numbers (RIN)< 2.0were excluded. Extracted RNA was stored at -80°C until downstream applications.

##### NGS Library prep and Sequencing

Libraries were prepared with the SMARTer Stranded Total RNAseq Kit v3 - Pico Input from 5 ng total RNA according to the manufacturer’s instruction and next-generation sequenced.

#### Alignment and statistical analysis

Raw sequencing reads were processed through a standardized RNA-seq analysis pipeline. Initial quality trimming was performed using Trimmomatic (v0.39), with adapter removal (ILLUMINACLIP), head cropping (HEADCROP:7), and minimum read length enforcement (MINLEN:35). Trimmed reads were aligned to the human reference genome (hg19) using STAR (v2.7.10a), with a maximum of three allowed multimapping locations per read (--outFilterMultimapNmax 3) and minimum alignment score and match length thresholds relative to read length (--outFilterScoreMinOverLread 0.66, --outFilterMatchNminOverLread 0.66). To retain only primary alignments, reads were filtered using samtools (v1.12) with flag -F 260. Duplicate reads were identified and removed using a custom AWK script.

Gene-level quantification was performed using the featureCounts function from the Rsubread package (v2.0.1), referencing a comprehensive GTF annotation file from REFSEQ (https://hgdownload.soe.ucsc.edu/goldenPath/hg19/bigZips/genes/hg19.refGene.gtf.gz) and requiring a minimum mapping quality score of 2 (minMQS = 2). Differential gene expression analysis was carried out using the DESeq2 package (v1.30.1) in R. Count data were normalized and modeled comparing AIH and non-AIH samples. Wald tests were used to identify differentially expressed genes, and significance was assessed using Benjamini–Hochberg adjusted p-values. The regularized log (rlog) transformation was applied to the normalized count matrix for visualization and clustering analyses.

### Statistical analysis

#### Differential genes compared to healthy

Statistical analysis of differential genes was performed as previously reported. Briefly, for every gene in every sample, we test whether the observed gene coverage is higher than expected according to the healthy mean/variance estimated from a control group of 1230 self-reported healthy donors. Using each sample’s background rate and the scaling factor accounting for sequencing depth, we define an expected distribution and estimate the probability of the observed coverage under the null hypothesis that the sample originated from a healthy population. Genes with a FDR-corrected P-value below 0.001 are reported as significantly elevated in the sample. We are aware that the healthy cohort used to define the baseline consists of healthy adults. However, testing a separate cohort of samples from healthy children, we observe virtually no difference (Fig S1), which justifies the use of this reference.

#### Cell type composition of samples (deconvolution)

To estimate the tissue composition of every sample, we used a non-negative least square model as implemented in the ‘*nnls’* R package (1.4). Given reference matrix *X*^∊*KxG*^ of the genes in *K* cell types and vector *Y*^∊*G*^ of observed gene counts in a sample, the objective is identifying non-negative coefficients β̂(cell-type proportion) by solving 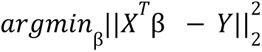 subject to β*_i_* ≥ 0 and 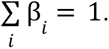 For reference tissue atlas we used 182 samples from the Roadmap and Blueprint H3K4me3 ChIP-seq data. Estimated coefficients of similar cell-types were summed and the final composition across 36 distinct cell-types is shown. These results were reproducible when using different features for the regression and with other regression models. A full list of cell-types used as reference data can be found in **Supplementary table 5**.

#### Liver single-cell signatures

Identification of liver specific cell-types genes was done based on liver specific marker genes from published liver scRNA-seq data(MacParland et al. 2018). To increase the specificity of cell-type signature in the cfDNA context, we exclude genes with mean counts above 1 in the healthy reference assuming their promoter is marked by H3K4me3 in non-liver cells contributing to the circulation. This filtering step resulted in a reduced number of marker genes particularly in the liver immune cells. A summary of the marker genes used for every liver cell-type can be found in **Supplementary Table 4**.

#### Expected, residual, and unexplained gene counts

For every sample we define the expected gene counts to be the mean gene counts of the composing cell types weighted by the contribution fraction of the cell-type as described above (*X* · β̂). To mitigate misleading results arising from missing tissues in the reference atlas, we added an additional healthy profile to the atlas, derived from a large cohort of healthy cfChIP-seq samples.

The residual is defined as *log*_2_ (1 + *observed*) − *log*_2_ (1 + *expected*). To further account for inter-tissue variability, we estimate the expected variance of every gene based on the weighted empirical variance observed in the replicates of the tissues composing the samples and test whether the null hypothesis that the observed counts are negative binomial distributed with that mean and variance can be rejected.

Formally, given a set of genes *G*, let:

*S*_1_, *S*_2_ … *S_n_* - set of cfChIP-seq samples

*Y_i,g_* - coverage of gene *g* in samples *i*

*B_i,g_* - background reads in gene *g* of sample *i*

*Q_i_* - normalization factor of sample *i* (sequencing rate)

*k* = 1,… *K* - set of reference cell-types

*X_k,g_* - coverage of gene *g* in cell-type *k*

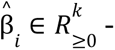 estimated fractions of cell-types composing sample *i*

*m_k,g_*, σ*_k,g_* - mean and standard deviation of gene *g* in cell-type *k*, were µ*_k,g_*, *m_k,g_* are estimated as previously described (Sadeh et al., 2021).

For every sample *i* the objective is to estimate the distribution:

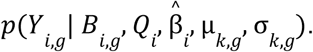

Due to discrete sampling in library preparation and sequencing we assume that *Y_i,g_* is Poisson distributed depending on the expected counts and sequencing depth.

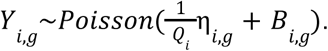

where η = *X* · β̂.

We approximate the distribution of *Y_i,g_* as negative binomial 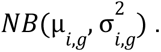 Using linearity of expectation and the law of total variation we can match the mean and variance of the negative binomial to that of the exact distribution:

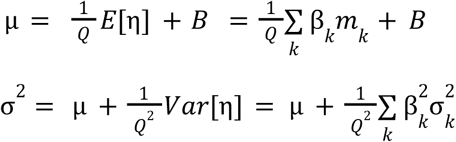

For every gene in every sample we compute the probability of *p*_NB_(*x* ≥ *Y_i,g_* |µ, σ^2^). Unexplained genes were defined as genes where the FDR-corrected q-value was less than 0.001 in at least 3 AIH samples.

#### AIH-specific regions and AIH-score

Samples from the AIH and non-AIH cohorts with elevated ALT levels (>40 U/L) were randomly divided into training, test, and validation sets. A two-sample *t*-test was performed on the residual signal between AIH and non-AIH samples in the training set to identify genomic regions that significantly deviate from the expected signal based on composition-informed reference with elevated signal in AIH. Regions with a Q-value < 0.1 were retained, after excluding those with elevated signal in healthy control samples. Subsequently, regions where the signal in the test set was less than 70% of the corresponding signal in the training set (i.e., test/train ratio < 0.7) were discarded.

The remaining regions were ranked by p-value, and a leave-one-patient-out cross-validation approach was used to determine the optimal number of regions to include in the classifier. The AIH-score was defined as the cumulative residual signal across the selected regions. Receiver Operating Characteristic (ROC) analysis was performed using the pROC R package.

#### PBC/PSC-specific regions and PBC/PSC-score

Differential signal analysis was performed by comparing the mean signal of PBC/PSC samples against this AIH reference group. Genomic regions were initially filtered for those showing a nominal p-value < 0.1 (two-sample t-test), a minimum signal intensity > 0.5 in the PBC/PSC cohort, and a fold-change > 1.2 relative to AIH. To ensure specificity, regions with high baseline signal in healthy controls (mean signal > 1) were excluded.

The top 40 regions ranked by fold-change were identified as PBC/PSC candidates. Conversely, regions significantly depleted in PBC/PSC relative to AIH (p-value < 0.05, fold-change < 0.9) were identified as hepatocyte-associated candidates. Following this automated pipeline, candidates were manually curated to exclude regions exhibiting high intra-group variability or residual signal in healthy individuals. The final PBC/PSC-score was calculated as the cumulative signal across these refined genomic regions.

## Data Availability

All data produced in the present study will be deposited in public repository. And will be available upon reasonable request.

## Acknowledgements

We would like to thank Ahmad Quteineh for his assistance with RNA extraction and Daphna Josef-Strauss, Abed Nasereddin, and Idit Shiff for their assistance with RNA library preparation and sequencing. We thank the patients and their families for participating in this study.

This work was supported by Israel Science Foundation IPMP grant #3751/21 (to NF and EG) and by European Research Council grants #101189389, POC AIHLiquidBiopsy, and #101019560, AdG cfChIP, (both to NF).

## Data availability

Plasma cfChIP-seq normalized promoter coverage is available in the Zenodo repository https://zenodo.org/records/15322324. UCSC Track hub with cfChIP tracks is available at https://files.cs.huji.ac.il/nirf/track_hubs/AIH-cfChIP/hub.txt.

## Code availability

All script files used in the analysis in this manuscript can be found at https://github.com/Nir-Friedman-Lab/AIH_cfChIP

## Supplementary information

- Supplementary Table 1 - Individuals and samples clinical information

- Supplementary Table 2 - Sequencing statistics for samples sequenced in this study

- Supplementary Table 3 - AIH elevated genes (Fig. 1D)

- Supplementary Table 4 - liver cell-type marker genes (Fig. 2C)

- Supplementary Table 5 - Reference Tissues and Cell-types used for Deconvolution

- Supplementary Table 6 - Genes Unexplained by Tissue Composition (Fig. 3C)

- Supplementary Table 7 - genomic regions that comprise the AIH signature

## Supplementary figures

**Figure S1:**
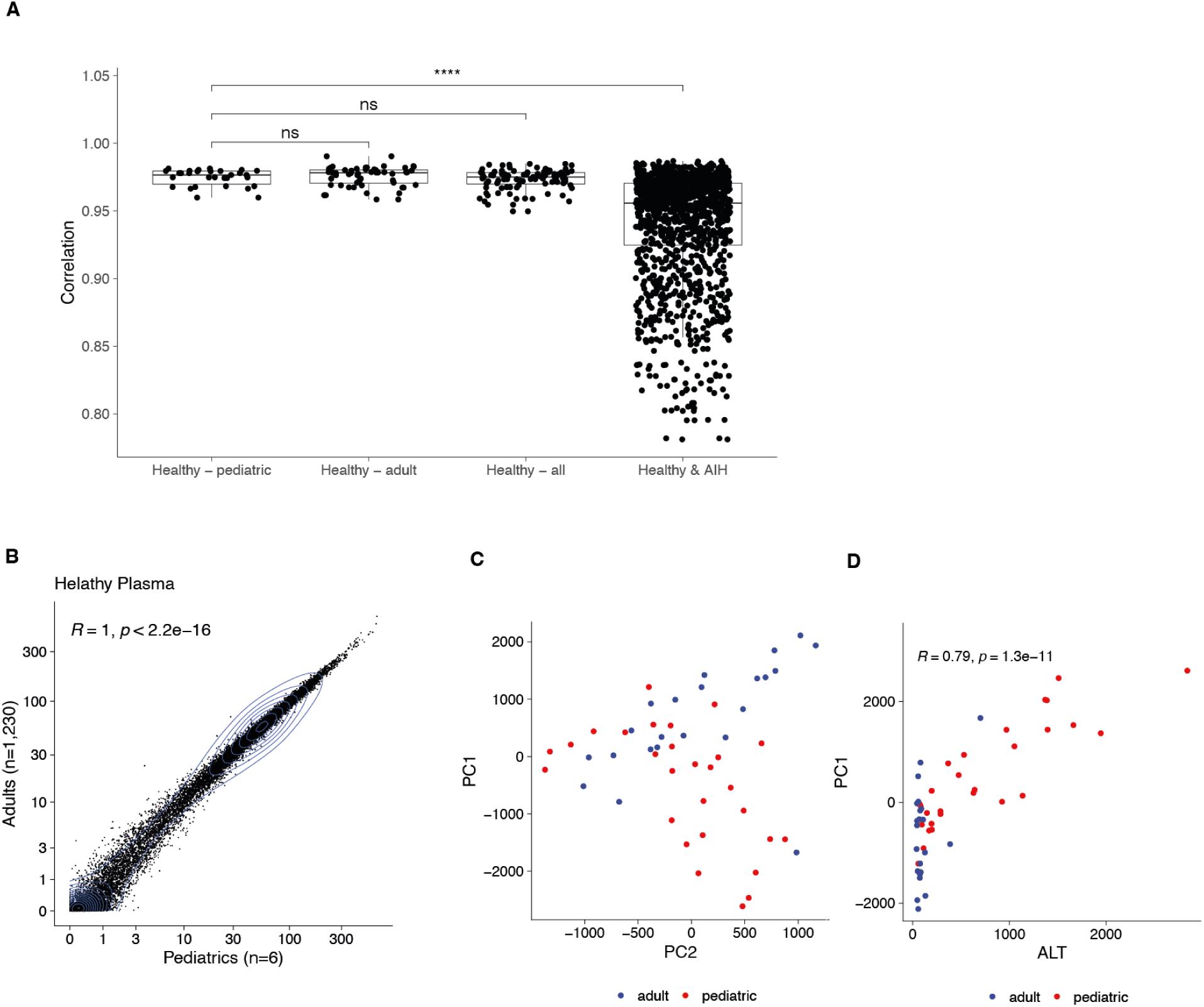
cfChIP-seq yield and healthy samples correlations. a. Promoter coverage pearson correlation (such as in panel B) within the healthy pediatric samples, healthy adults samples and between the adults vs. pediatrics healthy samples. The median correlation within these groups is not significant. Right most boxplot represents the same correlation comparing healthy vs. AIH samples. The median correlation of this cross-group comparison is significantly lower. b. Evaluation of similarity of plasma samples from healthy adults (n=1,230) and healthy pediatrics. For each gene, the mean normalized promoter coverage in healthy adult samples (y-axis) was compared to that of pediatric samples (x-axis). c. Principal components 1 and 2 AIH pediatric and adult samples. Principal component analysis was done using all Refseq genes (∼25,000). d. PC1 - the main source of variation among the AIH samples (∼50%), is correlated to ALT levels of the samples.

**Figures S2:**
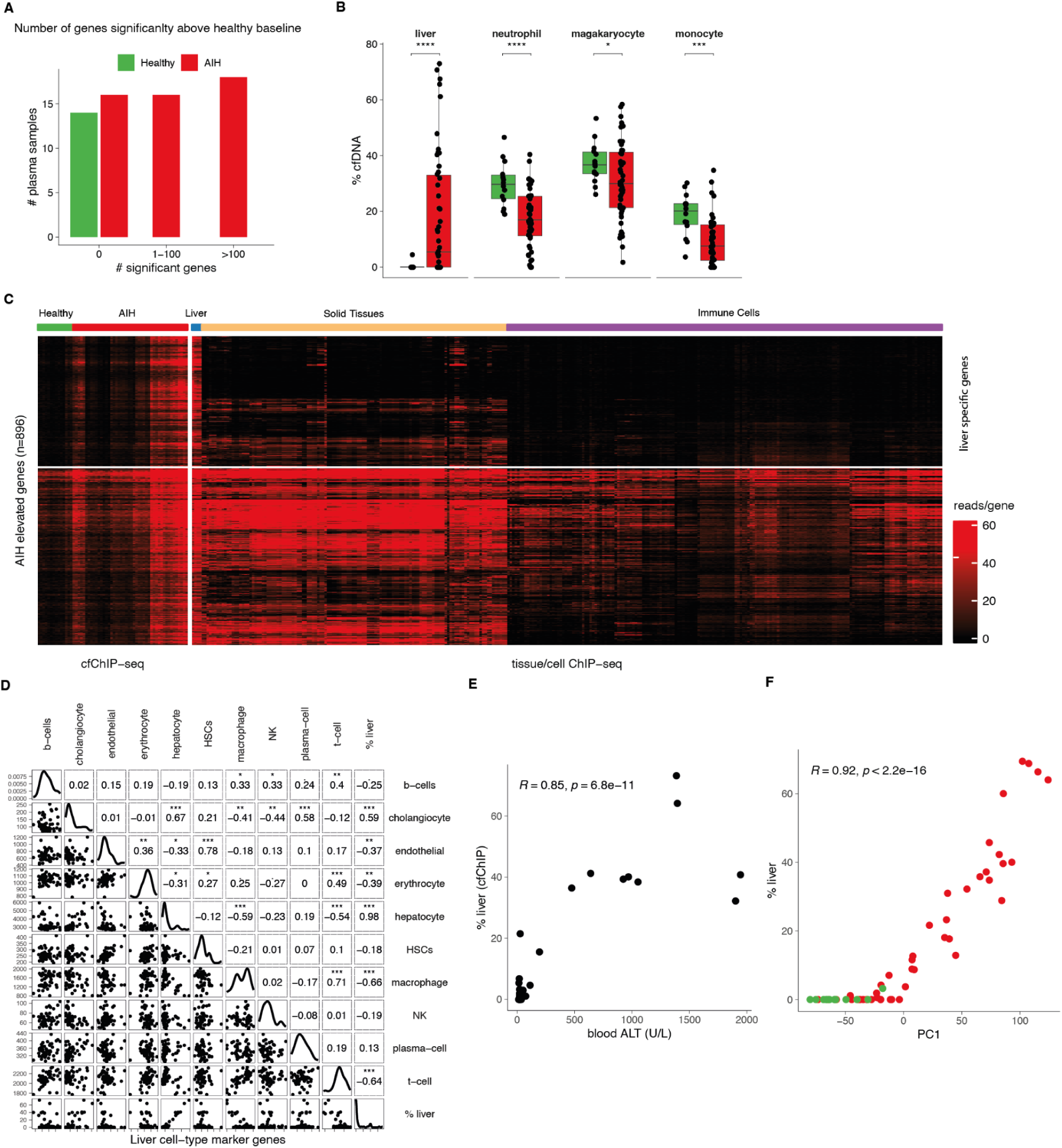
Deconvolution and transcriptional characterization of liver-derived cfDNA in AIH. a. **Differential gene burden.** Histogram of genes significantly elevated above healthy baseline in AIH (red) vs. healthy controls (green). AIH samples display a high frequency of differentially elevated genes, which are absent in healthy controls. b. **cfDNA cellular deconvolution.** Estimated relative fractions of liver and major hematopoietic lineages. AIH plasma shows a significant increase in liver-derived cfDNA and a relative decrease in the hematopoietic background compared to healthy donors. c. **Tissue specificity of AIH-elevated genes.** Hierarchical clustering of genes significantly elevated in at least three AIH samples (rows) across cfChIP-seq samples and a comprehensive reference atlas of 36 tissues and cell types (columns). The heat map demonstrates that genes elevated in AIH plasma are specifically enriched in liver reference samples and largely absent from immune cell types, which constitute the primary source of cfDNA in healthy individuals. d. **Cellular source validation.** Pearson correlation between the total liver deconvolution fraction and a cumulative hepatocyte-specific gene signature. The strong correlation identifies hepatocytes as the dominant source of liver cfDNA. e. **Clinical correlation.** Correlation between liver cfDNA fraction and serum ALT levels, demonstrating agreement with standard biochemical markers. f. **Drivers of transcriptomic variance.** Correlation between the first principal component (PC1) and estimated liver fraction, showing that hepatocellular DNA is the primary axis of variation in AIH plasma.

**Figure S3:**
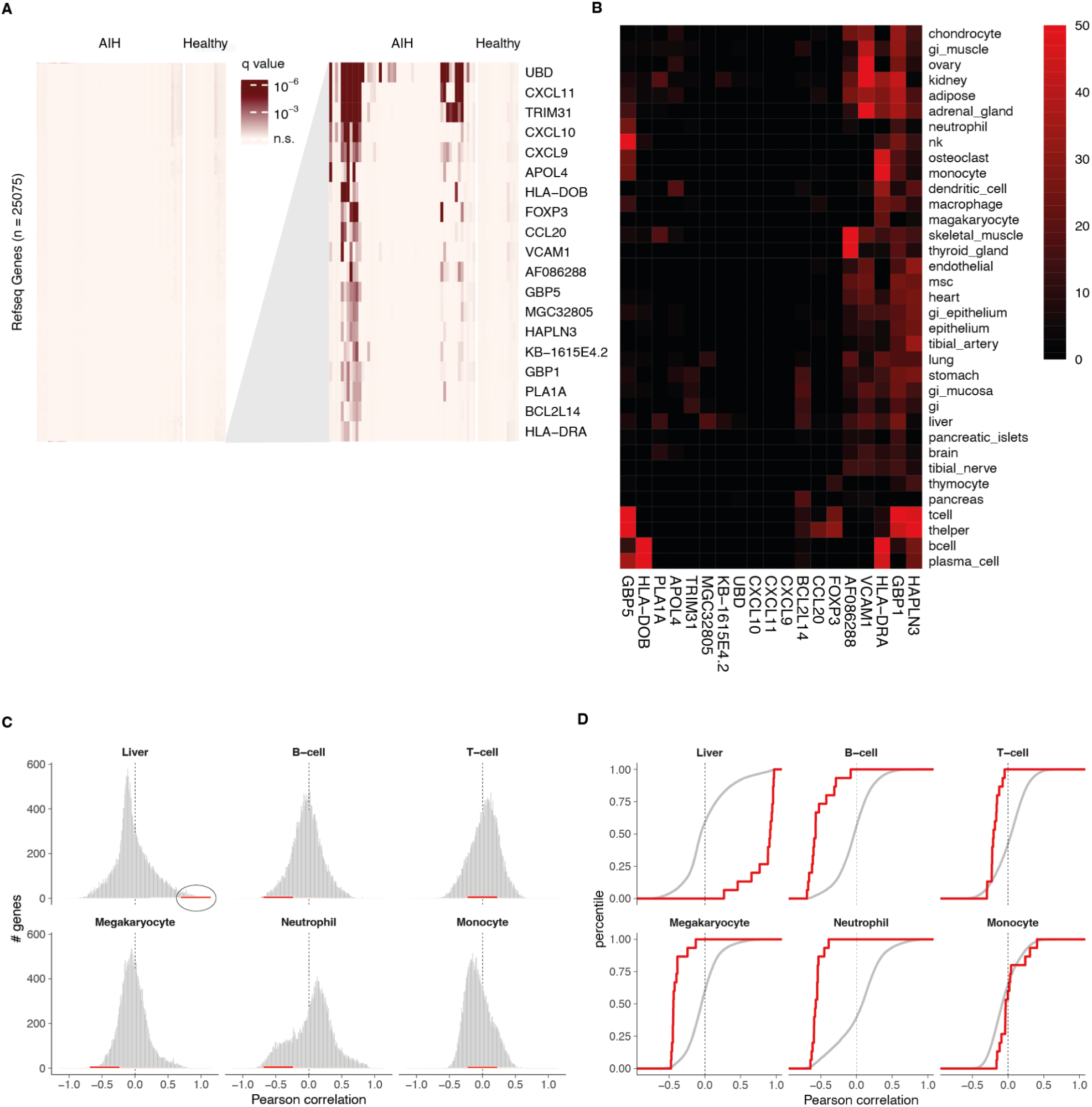
residual cf-ChIP-seq signal unexplained by tissue composition. a. Right: heatmap of all Refseq genes across AIH and Healthy samples. Color indicates the probability of the observed signal given the composition of the sample (methods). The majority of the genes don’t deviate significantly above expected. Left: zoom in a subset of genes with significantly elevated signal above expected by tissue composition in multiple AIH samples. b. Patterns of promoter coverage of reference tissues and cell types (rows) over genes with elevated signal unexplained by tissue composition (columns). Color represents normalized coverage over the promoter region of the genes. c. Correlation of promoter coverage and estimated tissue fraction in the AIH samples. Gray background represents the correlation of all Refseq genes, and the red histogram represents the correlation of the 11 unexplained genes shown in A. d. the cumulative distribution function of correlation shown in C. The black and red lines represent all Refseq genes and unexplained genes, respectively.

**Figure S4:**
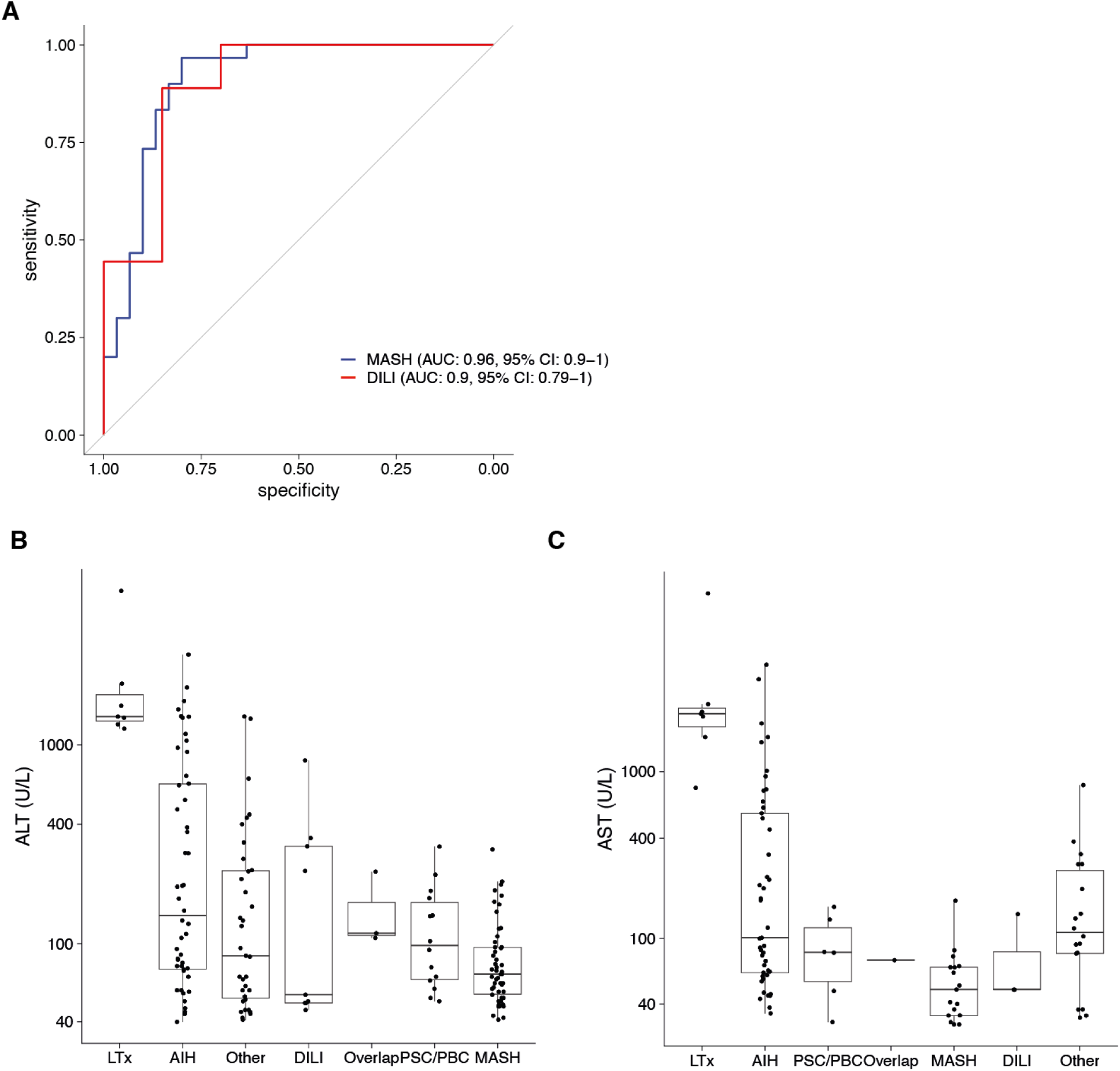
Liver enzymes levels poorly distinguish AIH from other liver diseases. A. Classifier performance for contrasting AIH vs MASH and DILI among samples with elevated transaminase levels. B. Boxplots of ALT across the various cohorts. ALT levels are particularly high in patients recovering liver transplant. Among the other groups, ALT is a poor predictor of AIH. C. Same for AST.

**Figure S5:**
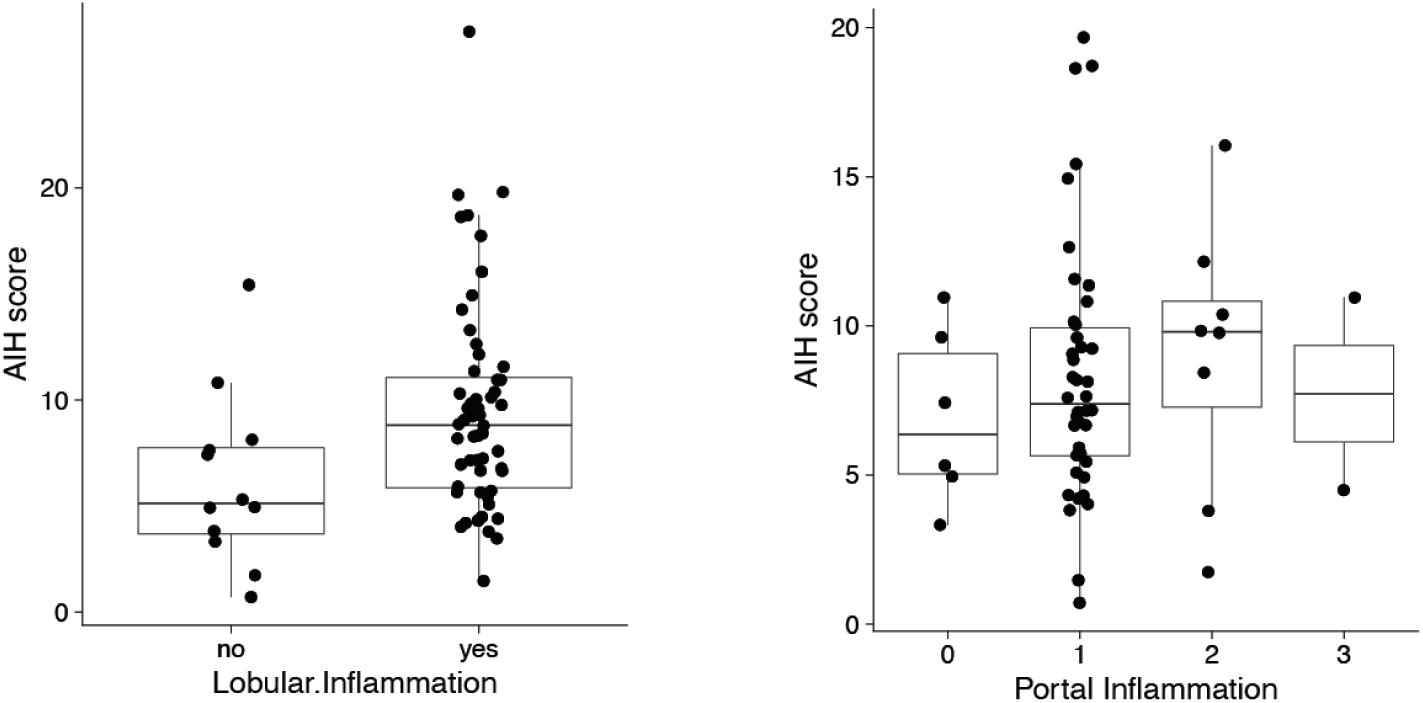
AIH score is low in MASH samples, including samples with marked inflammation.

**Figure S6:**
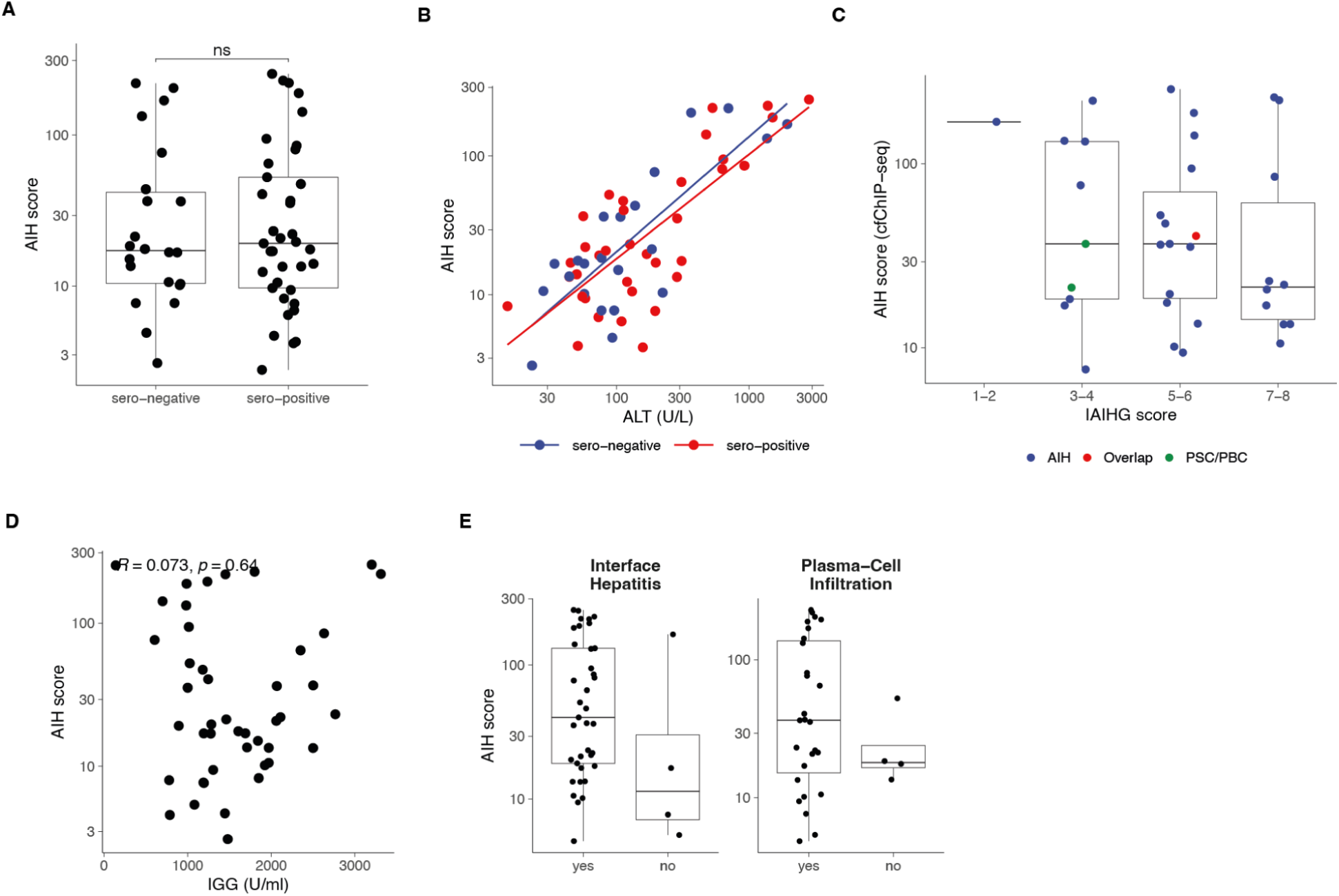
Association of the AIH score with serological, histological, and immunological features of autoimmune hepatitis. A. Distribution of the AIH score in seropositive and seronegative patients. No significant difference is observed between the two groups, indicating that the cfChIP-seq–derived score is largely independent of autoantibody status. B. Scatter plot showing the relationship between the AIH score and serum ALT levels, stratified by serological status. Trend lines demonstrate a similar association between ALT and the AIH score in both seropositive and seronegative patients. C. Comparison of the plasma based AIH-score (y-axis) and the International Autoimmune Hepatitis Group (IAIHG) score (x-axis). Only samples with sufficient clinical and laboratory data to calculate an IAIHG score are included. D. Comparison of the AIH score across histopathological features: interface hepatitis (left) and plasma cell infiltration (right). Samples with histological evidence of these features tend to show higher AIH scores, although differences do not reach statistical significance. E. Relationship between the AIH score and serum IgG levels. No significant correlation is observed, indicating that the cfChIP-seq–derived signal is largely independent of systemic immunoglobulin concentrations.

