## Supplementary methods for "Plasma cfChIP-seq for non-invasive identification of autoimmune liver diseases"

### Statistical analysis

#### Cell type composition of samples (deconvolution)

To estimate the tissue composition of every sample, we used a non-negative least square model as implemented in the '*nnls*' R package (1.4). Given reference matrix $X^{\in KxG}$ of the genes in $K$ cell types and vector $Y^{\in G}$ of observed gene counts in a sample, the objective is identifying non-negative coefficients $\hat{\beta}$ (cell-type proportion) by solving $argmin_{\beta}||X^T\beta - Y||_2^2$ subject to $\beta_i \geq 0$ and $\sum_i \beta_i = 1$. For reference tissue atlas we used 182 samples from the Roadmap and Blueprint H3K4me3 ChIP-seq data. Estimated coefficients of similar cell-types were summed and the final composition across 36 distinct cell-types is shown. These results were reproducible when using different features for the regression and with other regression models. A full list of cell-types used as reference data can be found in **Supplementary table 5** .

#### Liver single-cell signatures

Identification of liver specific cell-types genes was done based on liver specific marker genes from published liver scRNA-seq data[2]. To increase the specificity of cell-type signature in the cfDNA context, we exclude genes with mean counts above 1 in the healthy reference assuming their promoter is marked by H3K4me3 in non-liver cells contributing to the circulation. This filtering step resulted in a reduced number of marker genes particularly in the liver immune cells. A summary of the marker genes used for every liver cell-type can be found in **Supplementary Table 4**.

#### Expected, residual, and unexplained gene counts

For every sample we define the expected gene counts to be the mean gene counts of the composing cell types weighted by the contribution fraction of the cell-type as described above $(X \cdot \hat{\beta})$. To mitigate misleading results arising from missing tissues in the reference atlas, we added an additional healthy profile to the atlas, derived from a large cohort of healthy cfChIP-seq samples.

The residual is defined as $log_2(1 + observed) - log_2(1 + expected)$. To further account for inter-tissue variability, we estimate the expected variance of every gene based on the weighted empirical variance observed in the replicates of the tissues composing the samples and test whether the null hypothesis that the observed counts are negative binomial distributed with that mean and variance can be rejected.

Formally, given a set of genes $G$, let:

$S_1, S_2 ... S_n$ - set of cfChIP-seq samples

$Y_{i,g}$ - coverage of gene $g$ in samples $i$

$B_{i,g}$ - background reads in gene $g$ of sample $i$

$Q_i$ - normalization factor of sample $i$ (sequencing rate)

$k = 1, ... K$ - set of reference cell-types

$X_{k,g}$ - coverage of gene $g$ in cell-type $k$

$\hat{\beta}_i \in R^k_{\geq 0}$ - estimated fractions of cell-types composing sample $i$

$m_{k,g}, \sigma_{k,g}$ - mean and standard deviation of gene $g$ in cell-type $k$, were $\mu_{k,g}, m_{k,g}$ are estimated as previously described (Sadeh et al., 2021).

For every sample $i$ the objective is to estimate the distribution:

$$p(Y_{i,g} | B_{i,g}, Q_i, \hat{\beta}_i, \mu_{k,g}, \sigma_{k,g}).$$

Due to discrete sampling in library preparation and sequencing we assume that $Y_{i,g}$ is Poisson distributed depending on the expected counts and sequencing depth.

$$Y_{i,g} \sim Poisson(\frac{1}{Q_i}\eta_{i,g} + B_{i,g}).$$

where $\eta = X \cdot \hat{\beta}$.

We approximate the distribution of $Y_{i,g}$ as negative binomial $NB(\mu_{i,g}, \sigma^2_{i,g})$. Using linearity of expectation and the law of total variation we can match the mean and variance of the negative binomial to that of the exact distribution:

$$\mu = \frac{1}{Q}E[\eta] + B = \frac{1}{Q}\sum_k \beta_k m_k + B$$

$$\sigma^2 = \mu + \frac{1}{Q^2}Var[\eta] = \mu + \frac{1}{Q^2}\sum_k \beta_k^2 \sigma_k^2$$

For every gene in every sample we compute the probability of $p_{NB}(x \geq Y_{i,g} | \mu, \sigma^2)$. Unexplained genes were defined as genes where the FDR-corrected q-value was less than 0.001 in at least 3 AIH samples.

$k = 1,... K$ - set of reference cell-types

$X_{k,g}$ - coverage of gene $g$ in cell-type $k$

$\hat{\beta}_i \in R^k_{\geq 0}$ - estimated fractions of cell-types composing sample $i$

$m_{k,g}, \sigma_{k,g}$ - mean and standard deviation of gene $g$ in cell-type $k$, were $\mu_{k,g}, m_{k,g}$ are estimated as previously described (Sadeh et al., 2021).

For every sample $i$ the objective is to estimate the distribution:

$$p(Y_{i,g} | B_{i,g}, Q_i, \hat{\beta}_i, \mu_{k,g}, \sigma_{k,g}).$$

Due to discrete sampling in library preparation and sequencing we assume that $Y_{i,g}$ is Poisson distributed depending on the expected counts and sequencing depth.

$$Y_{i,g} \sim Poisson(\frac{1}{Q_i}\eta_{i,g} + B_{i,g}).$$

where $\eta = X \cdot \hat{\beta}$.

We approximate the distribution of $Y_{i,g}$ as negative binomial $NB(\mu_{i,g}, \sigma^2_{i,g})$. Using linearity of expectation and the law of total variation we can match the mean and variance of the negative binomial to that of the exact distribution:

$$\mu = \frac{1}{Q}E[\eta] + B = \frac{1}{Q}\sum_k \beta_k m_k + B$$

$$\sigma^2 = \mu + \frac{1}{Q^2}Var[\eta] = \mu + \frac{1}{Q^2}\sum_k \beta_k^2\sigma_k^2$$

For every gene in every sample we compute the probability of $p_{NB}(x \geq Y_{i,g}|\mu, \sigma^2)$. Unexplained genes were defined as genes where the FDR-corrected q-value was less than 0.001 in at least 3 AIH samples.
